# Development and evaluation of an algorithm to link mothers and infants in two US commercial healthcare claims databases for pharmacoepidemiology research

**DOI:** 10.1101/2022.12.13.22283418

**Authors:** James Weaver, Jill H. Hardin, Clair Blacketer, Alexis A. Krumme, Melanie H. Jacobson, Patrick B. Ryan

**Author notes:** Corresponding author James Weaver. **Source of funding** This work was conducted as part of employment at Johnson & Johnson. **Author contributions** JW: Methodology, Software, Formal analysis, Writing - Original Draft, Visualization; JHH: Methodology, Software, Writing - Original Draft; CB: Methodology, Software, Writing – Review & Editing; AAK: Writing - Review & Editing; MHJ: Writing - Review & Editing; PBR: Conceptualization, Methodology, Writing - Review & Editing. **Condensation** This research developed and evaluated an algorithm to link mothers and infants in two US commercial healthcare databases to facilitate research on prenatal medication exposure and infant health outcomes.

## Abstract

**Background:** Administrative healthcare claims databases are used in drug safety research but are limited for investigating the impacts of prenatal exposures on neonatal and pediatric outcomes without mother-infant pair identification.

**Objective:** We developed a mother-infant linkage algorithm and evaluated it in two, large US commercially insured populations.

**Study Design:** We used two US commercial health insurance claims databases during the years 2000 to 2021. Mother-infant links were constructed where persons of female sex 12-55 years of age with a pregnancy episode ending in live birth were associated with a person who was 0 years of age at database entry, who shared a common insurance plan ID, had overlapping insurance coverage time, and whose date of birth was within ±60-days of the mother’s pregnancy episode live birth date. We compared the characteristics of linked vs non-linked mothers and infants to assess similarity.

**Results:** The algorithm linked 3,477,960 mothers to 4,160,284 infants in the two databases. Linked mothers and linked infants comprised 73.6% of all mothers and 49.1% of all-infants, respectively. 94.9% of linked infants’ dates of birth were within ±30-days of the associated mother’s pregnancy episode end dates. Linked mothers were older, had longer pregnancy episodes, and had greater post-pregnancy observation time than mothers with live births who did not meet linkage algorithm criteria. Linked infants had less observation time and greater healthcare utilization than non-linked infants. Other characteristics were similar in linked vs non-linked mothers and infants.

**Conclusion:** We developed a mother-infant linkage algorithm and applied it to two US commercial healthcare claims databases that achieved a high linkage proportion and demonstrated that linked and non-linked mother and infant cohorts were similar. Transparent, reusable algorithms applied to large databases enables large-scale research on exposures during pregnancy and pediatric outcomes with relevance to drug safety. These features suggest that prenatal exposure causal risk assessment that uses this algorithm can produce valid and generalizable evidence to inform clinical, policy, and regulatory decisions.

**Key points:** A. Why was this study conducted?

-This study establishes reliable mother-infant links in two US commercial healthcare databases to facilitate research on prenatal exposures and infant health outcomes

B. What are the key findings?

-Linked mothers with live births comprise 73.6% of all mothers with live births and linked infants comprise 49.1% of all infants

-Linked vs. non-linked mother and infant cohorts have similar demographic and clinical profiles

-Substantial linked coverage and linked vs non-linked characteristic similarity suggests that prenatal exposure causal risk assessment using the linked cohorts will produce valid and generalizable evidence

C. What does this study add to what is already known?

-This study created large mother-infant linked cohorts to enable research on rare exposures and outcomes available in healthcare claims databases

-Linked mother and infant coverage is similar to that reported in previous linkage studies

-Descriptive comparisons between linked vs. non-linked mother and infant cohorts increases confidence that results from research on linked cohorts also apply to mother and infant populations that do not meet linkage algorithm criteria

-This mother-infant linkage algorithm is publicly available and easily implemented in databases converted to a common data model

## Introduction

Pregnancy is characterized by distinct periods of embryonic development representing critical exposure windows for children’s health [1]. Exposures before or during pregnancy, including pharmaceuticals, can affect conception, fetal development, pregnancy outcomes, and children’s health. While up to 90% of women take medication during pregnancy[2, 3], drug safety evidence is scarce because clinical trials often exclude pregnant people[4–6]. Mechanisms for generating pregnancy drug safety evidence are available, such as teratology information services[7], pregnancy and birth registries [8–12], case control studies[13], prospective cohort studies[14], and linked registry and prescription data resources[15]. However, these approaches often lack power to adequately assess rare exposures or outcomes, suffer from information biases, are slow to deliver results, may reflect selected populations, and are resource intensive. This research landscape produces an incomplete understanding of the benefits and risks of prenatal medication use and resultant birth outcomes. Further, the COVID-19 pandemic highlighted the need for timely, robust evidence on the benefit and risks of prenatal vaccine use.

Calls have been made to use real-world data (RWD) to study medication effects in pregnancy and are increasingly accepted by health authorities as part of post-authorization safety commitments[16, 17]. Large, administrative healthcare databases for pregnancy research are advantageous because they include large samples, multi-therapeutic area drug dispensing and diagnosis reimbursement claims, longitudinal patient observation, and reflect routine-care clinical practice.

To assess prenatal exposures on infant outcomes in RWD requires implementing algorithms to define pregnancy episodes and to link live births to infant records, which is challenging in the United States where national health record identifiers are absent. Mother-infant linkage has been conducted US administrative healthcare databases, including among Medicaid, commercially-insured, and Military Health System populations[18–23]. Other efforts, such as the Medication Exposure in Pregnancy Risk Evaluation Program (MEPREP)[24, 25], have linked administrative and electronic health record data to state birth records. However, details on linkage confidence and evaluation are sparse[26].

Our study builds on past efforts to create mother-infant linked cohorts in RWD. We created the largest known cohorts of mother-infant linked data using two large, US commercial insurance databases. In contrast to other linkage studies that use proprietary algorithms, our algorithm is publicly available. The algorithm was developed for use against the Observational Medical Outcomes Partnership (OMOP) Common Data Model (CDM) [27, 28], so it may be applicable to similar databases that have been standardized. Our linkage algorithm furthers earlier linkage work based on insurance enrollment ID matching only, by applying additional temporal criteria intended to increase linkage confidence. We evaluated the algorithm through comprehensive characterization comparisons between linked and non-linked mothers and infants.

## Materials and Methods

### Data sources

The study used two health insurance claims databases, IBM® Marketscan® Commercial Database (CCAE)[2000–2022] and Optum’s de-identified Clinformatics® Data Mart Database (Clinformatics®)[2000–2021]. Both contain de-identified, patient-level, encounter-based, longitudinal, employer-based US administrative health insurance claims records and include inpatient and outpatient diagnoses, procedures, and outpatient prescription dispensing records. Both databases use a unique insurance enrollment ID for identifying beneficiaries and their dependents under a single, primary insurance holder account. Both databases were transformed to the OMOP CDM, which provides a standardized representation of database structure and clinical content[29] to enable consistent analysis across disparate healthcare databases[30, 31].

### Linkage algorithm

The linkage algorithm relies on and is distinct from identifying pregnancy episodes and outcomes[32]. The pregnancy episodes algorithm was previously described, implemented, and validated in several administrative healthcare databases, including those in this study[32]. Briefly, it identifies pregnancy outcomes and estimates pregnancy start and end dates using a hierarchy of pregnancy markers, such as last menstrual period, amenorrhea, urine tests, ultrasounds, and delivery procedures in females aged 12-55 years.

### Step 1: Identify candidate mothers and infants

We first identified candidate mothers as females whose pregnancy episode(s) ended with live birth and occurred during a period of insurance enrollment.

Multiple periods of insurance enrollment were combined into a single observation period provided gaps between an enrollment period end and subsequent start date were ≤30 days. We identified candidate infants as persons whose year of birth was the same as their first observation period start year (i.e., were 0 years of age at observation period start) and had an insurance enrollment ID shared with a candidate mother. Candidate infants’ date of birth (DOB) was set as year, month, and day. Year of birth was available for all persons in both databases. Where month and day were unavailable, we inferred these components from observation period start month and day. Most day of birth values were set as 1 because insurance enrollment typically begins on the first day of a month.

### Step 2: Identify candidate mother-infant links

We identified candidate links between mothers and infants where they matched on insurance enrollment ID and the candidate infant’s DOB occurred during a candidate mother’s observation period.

### Step 3: Classify probable mother-infant links

We identified probable links between mothers and infants by restricting to those where the candidate infant’s DOB occurred within ±60 days of the candidate mother’s pregnancy episode end date. This correspondence window was varied in a sensitivity analysis (Appendix Section A).

### Step 4: Exclude ambiguous mother-infant links

In Step 2, we identified rare instances where multiple mothers could be associated with a single infant. These records were excluded from analysis.

### Cohorts used in algorithm evaluation

Nine cohorts were constructed to compare characteristics between linked vs non-linked mothers and infants. The index date refers to the temporal reference against which covariates were constructed.

1) Mothers linked to ≥1 infant indexed at pregnancy episode start

2) Mothers linked to ≥1 infant indexed at pregnancy episode end

3) Infants linked to a mother indexed at pregnancy episode end

4) Mothers not linked to an infant indexed at pregnancy episode start

5) Mothers not linked to an infant indexed at pregnancy episode end

6) Infants not linked to a mother indexed at inferred DOB

7) Candidate mothers indexed at pregnancy episode start

8) Candidate mothers indexed at pregnancy episode end

9) Candidate Infants indexed at DOB

### Characterization analyses

We characterized mother cohorts relative to each index date: once with covariates that reflect events observed during the year before or on the pregnancy episode start date, and again with covariates that reflect events observed during the year before or on the pregnancy episode end date. We characterized the infant cohorts with covariates that reflect events observed on or during the year after the pregnancy episode end date. See Appendix Section B for demographic and clinical covariate details.

We compared characteristics between linked vs. non-linked mothers and infants to evaluate differences between populations that did and did not meet linkage algorithm criteria. We made covariate comparisons by calculating the standardized mean difference (SMD) for each covariate in units of the pooled standard deviation, a metric uninfluenced by large sample sizes[33], and interpreted SMD values >0.1 as meaningfully different[34, 35].

## Results

All source code and an interactive application for viewing full results is available at https://data.ohdsi.org/MotherInfantLinkEval/.

**Figure 1** depicts step-by-step attrition of the linkage algorithm.

**Figure 1.**
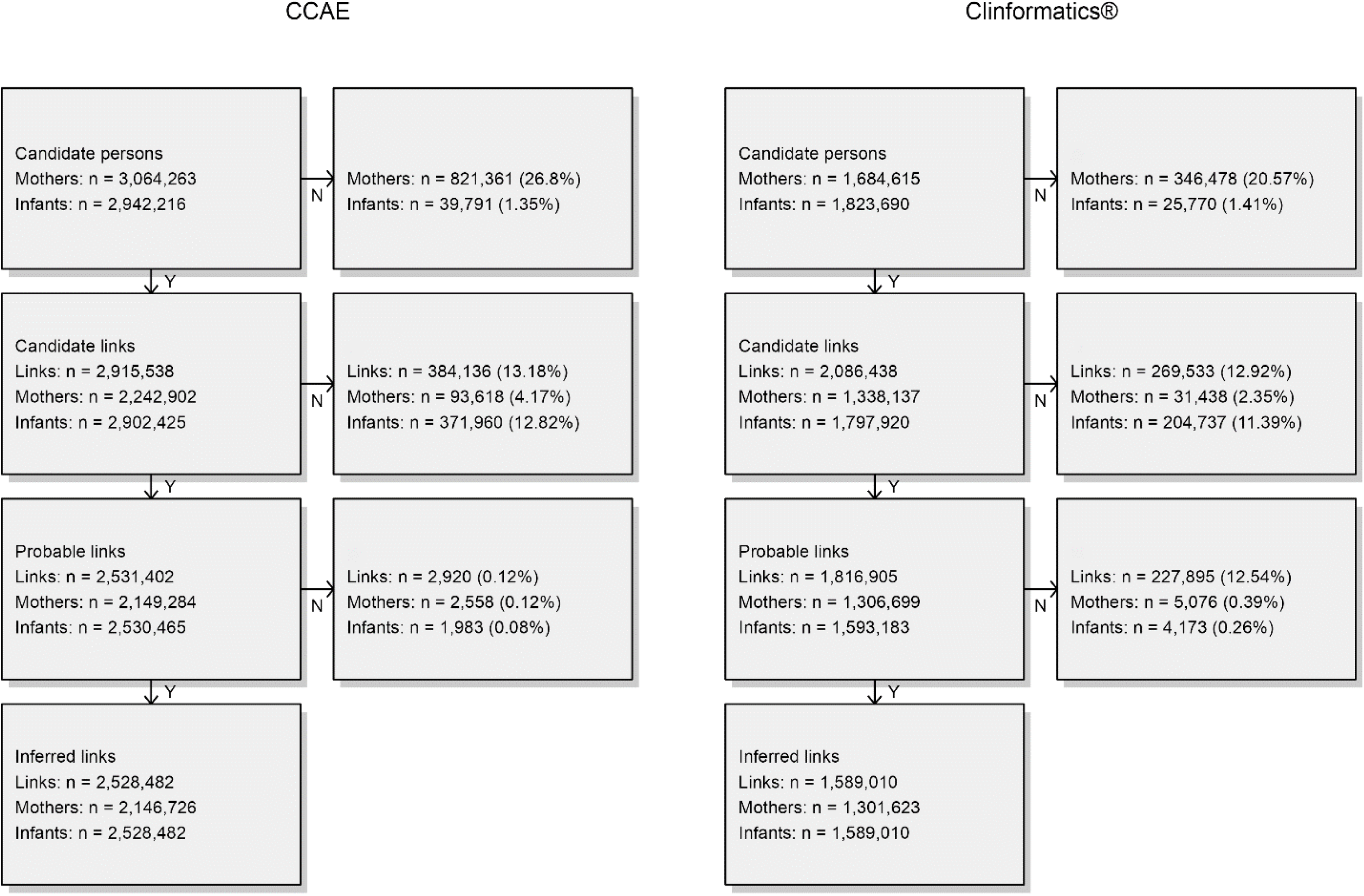
Mother-infant linkage algorithm attrition diagram. Primary algorithm implementation: first births, ±60-day pregnancy episode end/infant date-of-birth correspondence. CCAE: IBM® Marketscan® Commercial Database; Optum: Optum’s de-identified Clinformatics® Data Mart Database.

In CCAE, 3,064,263 candidate mothers and 2,942,216 candidate infants were identified in Step 1, of whom 26.8% and 1.4% were dropped respectively during Step 2, resulting in 2,915,538 candidate links. Links were reduced by 13.2% and 0.1% in steps 3 and 4 respectively, which resulted in 2,528,482 links: 2,146,726 linked mothers, and 2,528,482 linked infants. 31.3% of linked infant’s DOB were on the same day as their linked mother’s pregnancy episode end date and 58.3%, 71.5%, and 92.1% occurred within ±7 days, ±14 days, and ±30 days, respectively. Linked infant’s DOB was on average 5.9 days (SD=15.1, median=1) after the pregnancy episode end date. Linked mothers comprised 70.1% of all mothers (n=3,064,263) and linked infants comprised 51.2% of all infants (n=4,935,376)(Appendix Table C1).

In Clinformatics®, 1,684,615 candidate mothers and 1,823,690 candidate infants were identified, of whom 20.6% and 1.4% were dropped respectively during Step 2, resulting in 2,086,438 candidate links. Links were reduced by 12.9% and 12.5% in steps 3 and 4 respectively, which resulted in 1,589,010 links: 1,301,623 linked mothers and 1,589,010 linked infants. 67.4% of linked infant’s DOB were on the same day as their linked mother’s pregnancy episode end date and 98.0% 98.6%, and 99.3% occurred within ±7 days, ±14 days, and ±30 days, respectively. Linked infants’ DOB was on average 0.7 days (SD=4.0, median=0) after the pregnancy episode end date. Linked mothers comprised 77.3% of all mothers (n=1,684,615) and linked infants comprised 47.0% of all infants (n=3,379,811)(Appendix Table C1).

**Table 1** reports characteristics and SMDs of linked vs non-linked mothers for several characteristics measured relative to pregnancy episode start date. Pregnancy episode starts were equally distributed by year over the study period, although index dates in non-linked mothers were more common in February and March in CCAE. Mean age was greater among linked mothers in both databases (CCAE: 31.2 vs 27.4 years, Clinformatics®: 30.9 vs 27.9 years). There was greater post-pregnancy mean observation time among linked mothers in both databases (CCAE: 1358 vs 960 days, Clinformatics®: 1221 vs 930 days) and mean pregnancy episode length was greater among linked mothers in CCAE (273 vs 270 days). Linked vs non-linked mothers did not differ in clinical event counts, healthcare utilization, and health status in either database.

**Table 1.** Selected characteristics and standardized differences of linked and non-linked mothers in IBM® Marketscan® Commercial Database and Optum’s de-identified Clinformatics® Data Mart Database. Mother characteristics were measured on the pregnancy episode start date (index year and month, age) or during the 365-day period before and including the pregnancy episode start date (distinct event occurrence counts). Primary algorithm implementation: all births, ±60-day pregnancy episode end/infant date-of-birth correspondence.

| Characteristic | CCAЕ |  |  | Clinformatics® |  |  |
| --- | --- | --- | --- | --- | --- | --- |
|  | Linked<br>% (n =<br>2,528,482) | Non-linked<br>% (n =<br>995,892) | Std.Diff | Linked<br>% (n =<br>1,589,010) | Non-linked<br>% (n =<br>420,199) | Std.Diff |
| Index year |  |  |  |  |  |  |
| 2000 | 0.61 | 0.72 | 0.013 | 1.35 | 1.39 | 0.003 |
| 2001 | 0.94 | 1.13 | 0.019 | 3.83 | 3.77 | -0.003 |
| 2002 | 1.92 | 2.04 | 0.009 | 4.71 | 4.54 | -0.008 |
| 2003 | 3.11 | 2.64 | -0.028 | 4.84 | 4.65 | -0.009 |
| 2004 | 4.02 | 3.39 | -0.033 | 4.8 | 4.49 | -0.015 |
| 2005 | 3.97 | 3.27 | -0.037 | 6.18 | 4.09 | -0.095 |
| 2006 | 4.74 | 4.11 | -0.031 | 6.3 | 4.76 | -0.067 |
| 2007 | 4.93 | 4.36 | -0.027 | 6.31 | 4.81 | -0.065 |
| 2008 | 6.07 | 5.25 | -0.036 | 5.93 | 4.64 | -0.058 |
| 2009 | 6.17 | 5.83 | -0.014 | 5.12 | 5.18 | 0.003 |
| 2010 | 7.45 | 6.47 | -0.039 | 4.54 | 6.01 | 0.066 |
| 2011 | 8.16 | 8.36 | 0.007 | 4.37 | 6.45 | 0.092 |
| 2012 | 6.85 | 7.69 | 0.032 | 4.87 | 5.33 | 0.021 |
| 2013 | 6.96 | 7.26 | 0.012 | 4.71 | 4.81 | 0.005 |
| 2014 | 5.9 | 6.82 | 0.038 | 4.41 | 4.78 | 0.018 |
| 2015 | 5.34 | 5.99 | 0.028 | 4.6 | 5.02 | 0.02 |
| 2016 | 4.96 | 5.45 | 0.022 | 4.67 | 5.17 | 0.023 |
| 2017 | 4.87 | 5.11 | 0.011 | 4.81 | 5.28 | 0.022 |
| 2018 | 5.09 | 5.27 | 0.008 | 4.78 | 5.11 | 0.015 |
| 2019 | 4.5 | 4.79 | 0.014 | 4.44 | 4.89 | 0.021 |
| 2020 | 3.43 | 4.06 | 0.033 | 4.33 | 4.65 | 0.015 |
| 2021 | 0 | 0.01 | 0.008 | 0.11 | 0.18 | 0.017 |
| Index month |  |  |  |  |  |  |
| 1 | 10 | 9.71 | -0.01 | 9.05 | 9.14 | 0.003 |
| 2 | 7.58 | 10.71 | 0.109 | 7.79 | 8.32 | 0.019 |
| 3 | 6.1 | 14.36 | 0.275 | 7.86 | 9.19 | 0.048 |
| 4 | 7.4 | 8.38 | 0.037 | 7.42 | 7.7 | 0.011 |
| 5 | 8.48 | 7.07 | -0.053 | 8.24 | 7.97 | -0.01 |
| 6 | 8.42 | 6.8 | -0.061 | 8.07 | 7.8 | -0.01 |
| 7 | 8.88 | 6.89 | -0.074 | 8.41 | 8.11 | -0.011 |
| 8 | 8.79 | 6.84 | -0.073 | 8.37 | 8.01 | -0.013 |
| 9 | 8.7 | 7.1 | -0.059 | 8.52 | 8.18 | -0.012 |
| 10 | 8.73 | 7.43 | -0.048 | 8.79 | 8.48 | -0.011 |
| 11 | 8.49 | 7.23 | -0.047 | 8.7 | 8.4 | -0.011 |
| 12 | 8.43 | 7.47 | -0.036 | 8.78 | 8.71 | -0.003 |
| Age (years) |  |  |  |  |  |  |
| Mean | 31.22 | 27.36 | -0.483 | 30.93 | 27.94 | -0.357 |
| Std. deviation | 4.68 | 6.48 |  | 4.88 | 6.81 |  |
| Median | 31 | 27 |  | 31 | 28 |  |
| Prior observation time (days) |  |  |  |  |  |  |
| Mean | 737.21 | 833.74 | 0.079 | 715.62 | 778.33 | 0.054 |
| Std. deviation | 743.73 | 978.46 |  | 741.25 | 893.25 |  |
| Median | 503 | 489 |  | 476 | 472 |  |
| Post observation time (days) |  |  |  |  |  |  |
| Mean | 1357.52 | 959.98 | -0.265 | 1221.15 | 930.43 | -0.212 |
| Std. deviation | 1193.69 | 905.81 |  | 1092.51 | 829.61 |  |
| Median | 929 | 648 |  | 825 | 642 |  |
| Pregnancy episode length<br>(days) |  |  |  |  |  |  |
| Mean | 273.15 | 269.83 | -0.111 | 272.73 | 270.33 | -0.08 |
| Std. deviation | 18.44 | 23.58 |  | 18.89 | 23.27 |  |
| Median | 278 | 277 |  | 278 | 278 |  |
| Distinct conditions |  |  |  |  |  |  |
| Mean | 6.67 | 6.77 | 0.01 | 6.95 | 7.45 | 0.045 |
| Std. deviation | 6.73 | 7.34 |  | 7.38 | 8.18 |  |
| Median | 5 | 4 |  | 5 | 5 |  |
| Distinct drug ingredients |  |  |  |  |  |  |
| Mean | 4.68 | 4.62 | -0.009 | 4.14 | 4.44 | 0.04 |
| Std. deviation | 5.39 | 5.49 |  | 5.17 | 5.5 |  |
| Median | 3 | 3 |  | 3 | 3 |  |
| Distinct procedures |  |  |  |  |  |  |
| Mean | 7.4 | 6.62 | -0.075 | 7.08 | 6.92 | -0.015 |
| Std. deviation | 7.39 | 7.28 |  | 7.61 | 7.71 |  |
| Median | 5 | 5 |  | 5 | 5 |  |
| Distinct measurements |  |  |  |  |  |  |
| Mean | 6.7 | 6.29 | -0.025 | 13.62 | 12.48 | -0.03 |
| Std. deviation | 11.93 | 11.86 |  | 27.48 | 25.9 |  |
| Median | 3 | 3 |  | 3 | 3 |  |
| Distinct visit types |  |  |  |  |  |  |
| Outpatient Visit |  |  |  |  |  |  |
| Mean | 6.61 | 5.51 | -0.085 | 6.38 | 5.89 | -0.036 |
| Std. deviation | 9.81 | 8.31 |  | 10.47 | 8.98 |  |
| Median | 4 | 3 |  | 3 | 3 |  |
| Inpatient Visit |  |  |  |  |  |  |
| Mean | 0.11 | 0.11 | 0 | 0.1 | 0.11 | 0.003 |
| Std. deviation | 1.11 | 1.25 |  | 0.77 | 0.94 |  |
| Median | 0 | 0 |  | 0 | 0 |  |
| Emergency Room Visit |  |  |  |  |  |  |
| Mean | 0.16 | 0.34 | 0.09 | 0.38 | 0.33 | -0.007 |
| Std. deviation | 1.16 | 1.56 |  | 6.14 | 4.89 |  |
| Median | 0 | 0 |  | 0 | 0 |  |
| Charlson comorbidity index |  |  |  |  |  |  |
| Mean | 0.19 | 0.2 | 0.002 | 0.22 | 0.24 | 0.017 |
| Std. deviation | 0.85 | 0.89 |  | 0.93 | 1.02 |  |
| Median | 0 | 0 |  | 0 | 0 |  |

CCAE: IBM® MarketScan® Commercial Database; Clinformatics®: Optum's de-identified

**Table 2** reports characteristics and SMDs of linked vs non-linked mothers for the same characteristics as Table 1 except for pregnancy episode length but were measured relative to pregnancy episode end date. Age was greater among linked mothers in CCAE (32.0 vs 30.9 years), which reflects the slightly greater linked pregnancy episode lengths reported above. There was greater post-pregnancy observation time among linked mothers in both databases (CCAE: 1084 vs 690 days, Clinformatics®: 948 vs 660 days). Although uncommon, emergency room visits were greater among non-linked mothers in CCAE (0.7 vs 0.3).

**Table 2.** Selected characteristics and standardized differences of linked and non-linked mothers in IBM® Marketscan® Commercial Database and Optum’s de-identified Clinformatics® Data Mart Database. Mother characteristics were measured on the pregnancy episode end date (index year and month, age) or during the 365-day period before and including the pregnancy episode end date (distinct event occurrence counts). Primary algorithm implementation: all births, ±60-day pregnancy episode end/infant date-of-birth correspondence.

| Characteristic | CCAЕ |  |  | Clinformatics® |  |  |
| --- | --- | --- | --- | --- | --- | --- |
|  | Linked<br>% (n =<br>2,528,482) | Non-linked<br>% (n =<br>995,892) | Std.Diff | Linked<br>% (n =<br>1,589,010) | Non-linked<br>% (n =<br>420,199) | Std.Diff |
| Index year |  |  |  |  |  |  |
| 2000 | 0.1 | 0.31 | 0.047 | 0.01 | 0.02 | 0.01 |
| 2001 | 0.64 | 0.87 | 0.026 | 2.01 | 2.2 | 0.013 |
| 2002 | 1.14 | 1.51 | 0.032 | 4.27 | 4.29 | 0.001 |
| 2003 | 2.21 | 2.26 | 0.003 | 4.82 | 4.67 | -0.007 |
| 2004 | 3.32 | 2.87 | -0.026 | 4.78 | 4.57 | -0.01 |
| 2005 | 4.08 | 3.44 | -0.034 | 5.07 | 4.05 | -0.049 |
| 2006 | 4.03 | 3.37 | -0.035 | 6.37 | 4.42 | -0.087 |
| 2007 | 4.81 | 4.21 | -0.029 | 6.3 | 4.84 | -0.064 |
| 2008 | 5.26 | 4.71 | -0.025 | 6.26 | 4.79 | -0.064 |
| 2009 | 6.38 | 5.4 | -0.042 | 5.81 | 4.58 | -0.055 |
| 2010 | 6.17 | 6.26 | 0.004 | 4.8 | 5.62 | 0.037 |
| 2011 | 7.65 | 6.81 | -0.033 | 4.49 | 6.14 | 0.074 |
| 2012 | 8.04 | 8.94 | 0.032 | 4.4 | 6.46 | 0.091 |
| 2013 | 6.7 | 6.97 | 0.01 | 5.07 | 4.9 | -0.008 |
| 2014 | 6.97 | 7.38 | 0.016 | 4.56 | 4.74 | 0.009 |
| 2015 | 5.45 | 6.15 | 0.03 | 4.47 | 4.86 | 0.018 |
| 2016 | 5.44 | 6.03 | 0.025 | 4.73 | 5.12 | 0.018 |
| 2017 | 4.95 | 5.25 | 0.014 | 4.68 | 5.2 | 0.024 |
| 2018 | 4.91 | 5.18 | 0.012 | 4.8 | 5.23 | 0.02 |
| 2019 | 4.97 | 5.12 | 0.007 | 4.74 | 5.1 | 0.017 |
| 2020 | 4.43 | 4.68 | 0.012 | 4.34 | 4.76 | 0.021 |
| 2021 | 2.33 | 2.28 | -0.003 | 3.23 | 3.43 | 0.011 |
| Index month |  |  |  |  |  |  |
| 1 | 8.08 | 6.7 | -0.053 | 7.55 | 7.44 | -0.004 |
| 2 | 7.77 | 6.19 | -0.062 | 7.43 | 7.04 | -0.015 |
| 3 | 8.74 | 6.84 | -0.071 | 8.32 | 8.01 | -0.011 |
| 4 | 8.56 | 6.65 | -0.072 | 8.14 | 7.83 | -0.012 |
| 5 | 8.97 | 6.9 | -0.077 | 8.58 | 8.13 | -0.016 |
| 6 | 8.7 | 7.2 | -0.055 | 8.47 | 8.19 | -0.01 |
| 7 | 8.71 | 7.48 | -0.045 | 8.76 | 8.51 | -0.009 |
| 8 | 8.73 | 7.46 | -0.047 | 8.96 | 8.74 | -0.008 |
| 9 | 8.52 | 7.6 | -0.034 | 8.71 | 8.63 | -0.003 |
| 10 | 9.68 | 8.96 | -0.025 | 8.77 | 8.85 | 0.003 |
| 11 | 8.37 | 10.81 | 0.083 | 8.27 | 8.64 | 0.013 |
| 12 | 5.17 | 17.22 | 0.389 | 8.04 | 9.99 | 0.068 |
| Age (years) |  |  |  |  |  |  |
| Mean | 31.99 | 27.98 | -0.504 | 31.68 | 28.66 | -0.36 |
| Std. deviation | 4.67 | 6.44 |  | 4.87 | 6.81 |  |
| Median | 32 | 27 |  | 32 | 28 |  |
| Prior observation time (days) |  |  |  |  |  |  |
| Mean | 1010.36 | 1103.57 | 0.076 | 988.36 | 1048.67 | 0.052 |
| Std. deviation | 743.4 | 978.27 |  | 741 | 893.1 |  |
| Median | 777 | 758 |  | 749 | 743 |  |
| Post observation time (days) |  |  |  |  |  |  |
| Mean | 1084.37 | 690.15 | -0.263 | 948.41 | 660.09 | -0.21 |
| Std. deviation | 1192.66 | 905.33 |  | 1091.46 | 828.86 |  |
| Median | 656 | 377 |  | 552 | 371 |  |
| Distinct conditions |  |  |  |  |  |  |
| Mean | 19.81 | 20.75 | 0.061 | 20.21 | 21.43 | 0.076 |
| Std. deviation | 10.21 | 11.4 |  | 10.64 | 11.96 |  |
| Median | 18 | 18 |  | 18 | 19 |  |
| Distinct drug ingredients |  |  |  |  |  |  |
| Mean | 5.3 | 5.65 | 0.045 | 5.4 | 6 | 0.074 |
| Std. deviation | 5.35 | 5.61 |  | 5.39 | 5.84 |  |
| Median | 4 | 4 |  | 4 | 4 |  |
| Distinct procedures |  |  |  |  |  |  |
| Mean | 17.46 | 16.82 | -0.054 | 18.6 | 18.55 | -0.004 |
| Std. deviation | 8.44 | 8.51 |  | 8.72 | 9.11 |  |
| Median | 16 | 15 |  | 18 | 17 |  |
| Distinct measurements |  |  |  |  |  |  |
| Mean | 20.85 | 20.95 | 0.004 | 44.01 | 41.65 | -0.046 |
| Std. deviation | 15.57 | 16.57 |  | 36.44 | 36.03 |  |
| Median | 18 | 18 |  | 32 | 27 |  |
| Distinct visit types |  |  |  |  |  |  |
| Outpatient Visit |  |  |  |  |  |  |
| Mean | 15.53 | 14.21 | -0.089 | 15.7 | 14.76 | -0.054 |
| Std. deviation | 11.33 | 9.52 |  | 13.44 | 10.96 |  |
| Median | 13 | 12 |  | 12 | 12 |  |
| Inpatient Visit |  |  |  |  |  |  |
| Mean | 1.13 | 1.14 | 0.006 | 1.14 | 1.12 | -0.019 |
| Std. deviation | 0.67 | 0.69 |  | 0.71 | 0.64 |  |
| Median | 1 | 1 |  | 1 | 1 |  |
| Emergency Room Visit |  |  |  |  |  |  |
| Mean | 0.33 | 0.67 | 0.146 | 1.03 | 0.98 | -0.007 |
| Std. deviation | 1.36 | 1.85 |  | 5.63 | 4.52 |  |
| Median | 0 | 0 |  | 0 | 0 |  |
| Charlson comorbidity index |  |  |  |  |  |  |
| Mean | 0.3 | 0.3 | 0.001 | 0.32 | 0.34 | 0.016 |
| Std. deviation | 0.9 | 0.94 |  | 0.98 | 1.08 |  |
| Median | 0 | 0 |  | 0 | 0 |  |

CCAE: IBM Commercial Database; Clinformatics®: Optum's de-identified Clinformatics® Data

**Table 3** reports characteristics and SMDs of linked vs non-linked infants for several characteristics measured at the pregnancy episode end date. Non-linked births were more common in the early study period (2000-2002) in both databases and in CCAE linked births were more common in January. There was greater post-birth observation time among non-linked infants in both databases (CCAE: 886 vs 454 days, Clinformatics®: 751 vs 601 days). Condition (CCAE: 6.7 vs 5.7, Clinformatics®: 7.7 vs 6.4) and procedure (CCAE: 12.8 vs 9.9, Clinformatics®: 12.3 vs 10.0) occurrences were greater among linked infants. Healthcare utilization (i.e., outpatient and inpatient visits) was similarly greater among linked infants.

**Table 3.** Selected characteristics and standardized differences of linked and non-linked infants in IBM® Marketscan® Commercial Database and Optum’s de-identified Clinformatics® Data Mart Database. Infant characteristics were measured on the pregnancy episode end date (index year and month, age, sex) or during the 365-day period after and including the pregnancy episode end date (distinct event occurrence counts). Primary algorithm implementation: all births, ±60-day pregnancy episode end/infant date-of-birth correspondence.

| Characteristic | CCAЕ |  |  | Clinformatics® |  |  |
| --- | --- | --- | --- | --- | --- | --- |
|  | Linked<br>% (n =<br>2,528,482) | Non-linked<br>% (n =<br>2,406,894) | Std.Diff | Linked<br>% (n =<br>1,589,010) | Non-linked<br>% (n =<br>1,790,801) | Std.Diff |
| Sex |  |  |  |  |  |  |
| gender = FEMALE | 48.71 | 48.66 | -0.001 | 48.57 | 48.55 | 0 |
| gender = MALE | 51.29 | 51.34 | 0.001 | 51.43 | 51.45 | 0 |
| Index year |  |  |  |  |  |  |
| 2000 | 0.1 | 1.08 | 0.129 | 0.01 | 3.87 | 0.283 |
| 2001 | 0.64 | 1.07 | 0.046 | 2.01 | 6.89 | 0.238 |
| 2002 | 1.14 | 2.54 | 0.104 | 4.27 | 4.93 | 0.032 |
| 2003 | 2.21 | 3.95 | 0.1 | 4.82 | 4.66 | -0.008 |
| 2004 | 3.32 | 4.3 | 0.051 | 4.78 | 4.49 | -0.014 |
| 2005 | 4.08 | 4.44 | 0.018 | 5.07 | 6.65 | 0.067 |
| 2006 | 4.03 | 4.99 | 0.047 | 6.37 | 6.06 | -0.013 |
| 2007 | 4.81 | 4.8 | 0 | 6.3 | 5.97 | -0.014 |
| 2008 | 5.26 | 6.23 | 0.042 | 6.26 | 5.06 | -0.052 |
| 2009 | 6.38 | 6.57 | 0.008 | 5.81 | 4.13 | -0.077 |
| 2010 | 6.17 | 7.5 | 0.053 | 4.8 | 3.56 | -0.062 |
| 2011 | 7.65 | 7.35 | -0.011 | 4.49 | 3.52 | -0.05 |
| 2012 | 8.04 | 7.25 | -0.03 | 4.4 | 3.51 | -0.045 |
| 2013 | 6.7 | 6.32 | -0.015 | 5.07 | 3.99 | -0.052 |
| 2014 | 6.97 | 6.29 | -0.027 | 4.56 | 4.02 | -0.026 |
| 2015 | 5.45 | 4.15 | -0.061 | 4.47 | 4.68 | 0.01 |
| 2016 | 5.44 | 4.1 | -0.063 | 4.73 | 4.84 | 0.005 |
| 2017 | 4.95 | 4.02 | -0.045 | 4.68 | 4.68 | 0 |
| 2018 | 4.91 | 4.47 | -0.021 | 4.8 | 4.44 | -0.017 |
| 2019 | 4.97 | 3.65 | -0.065 | 4.74 | 4.04 | -0.034 |
| 2020 | 4.43 | 3.43 | -0.051 | 4.34 | 3.49 | -0.044 |
| 2021 | 2.33 | 1.49 | -0.062 | 3.23 | 2.54 | -0.041 |
| Index month |  |  |  |  |  |  |
| 1 | 8.08 | 4.57 | -0.145 | 7.55 | 6.15 | -0.055 |
| 2 | 7.77 | 6.37 | -0.055 | 7.43 | 5.91 | -0.061 |
| 3 | 8.74 | 7.42 | -0.048 | 8.32 | 6.73 | -0.06 |
| 4 | 8.56 | 8.49 | -0.003 | 8.14 | 6.97 | -0.045 |
| 5 | 8.97 | 8.58 | -0.014 | 8.58 | 10.15 | 0.054 |
| 6 | 8.7 | 9.04 | 0.012 | 8.47 | 8.5 | 0.001 |
| 7 | 8.71 | 10.52 | 0.062 | 8.76 | 9.93 | 0.04 |
| 8 | 8.73 | 9.88 | 0.039 | 8.96 | 9.57 | 0.021 |
| 9 | 8.52 | 10.31 | 0.061 | 8.71 | 9.95 | 0.042 |
| 10 | 9.68 | 9.67 | 0 | 8.77 | 9.38 | 0.021 |
| 11 | 8.37 | 7.88 | -0.018 | 8.27 | 8.32 | 0.002 |
| 12 | 5.17 | 7.28 | 0.087 | 8.04 | 8.44 | 0.014 |
| Post observation time<br>(days) |  |  |  |  |  |  |
| Mean | 454.64 | 886.47 | 0.269 | 601.17 | 751.11 | 0.1 |
| Std. deviation | 1154.08 | 1113.97 |  | 1127.43 | 985.63 |  |
| Median | 0 | 486 |  | 119 | 395 |  |
| Distinct conditions |  |  |  |  |  |  |
| Mean | 6.73 | 5.73 | -0.107 | 7.74 | 6.39 | -0.129 |
| Std. deviation | 6.88 | 6.39 |  | 7.63 | 7.11 |  |
| Median | 5 | 4 |  | 6 | 5 |  |
| Distinct drug ingredients |  |  |  |  |  |  |
| Mean | 2.64 | 2.45 | -0.038 | 8.53 | 7.44 | -0.065 |
| Std. deviation | 3.39 | 3.38 |  | 12.03 | 11.76 |  |
| Median | 1 | 1 |  | 2 | 2 |  |
| Procedures |  |  |  |  |  |  |
| Mean | 12.77 | 9.87 | -0.285 | 12.26 | 9.99 | -0.211 |
| Std. deviation | 7.38 | 7 |  | 7.82 | 7.36 |  |
| Median | 12 | 9 |  | 11 | 9 |  |
| Distinct measurements |  |  |  |  |  |  |
| Mean | 2.86 | 2.23 | -0.088 | 3.28 | 2.83 | -0.042 |
| Std. deviation | 5.06 | 4.92 |  | 7.41 | 7.61 |  |
| Median | 1 | 1 |  | 1 | 1 |  |
| Distinct visit types |  |  |  |  |  |  |
| Outpatient Visit |  |  |  |  |  |  |
| Mean | 10.36 | 8.65 | -0.185 | 9.45 | 8.32 | -0.105 |
| Std. deviation | 6.6 | 6.46 |  | 7.71 | 7.51 |  |
| Median | 9 | 8 |  | 8 | 7 |  |
| Inpatient Visit |  |  |  |  |  |  |
| Mean | 0.98 | 0.5 | -0.659 | 1.03 | 0.69 | -0.493 |
| Std. deviation | 0.47 | 0.57 |  | 0.47 | 0.51 |  |
| Median | 1 | 0 |  | 1 | 1 |  |
| Emergency Room Visit |  |  |  |  |  |  |
| Mean | 0.22 | 0.24 | 0.016 | 0.1 | 0.13 | 0.006 |
| Std. deviation | 0.84 | 0.91 |  | 3.94 | 3.45 |  |
| Median | 0 | 0 |  | 0 | 0 |  |

CCAE: IBM Commercial Database; Clinformatics®: Optum's de-identified Clinformatics® Data

**Figure 2** depicts the comparative prevalence of demographic, drug exposure, condition, procedure, and measurement occurrence covariates for the linked vs non-linked mother and infant cohorts.

**Figure 2.**
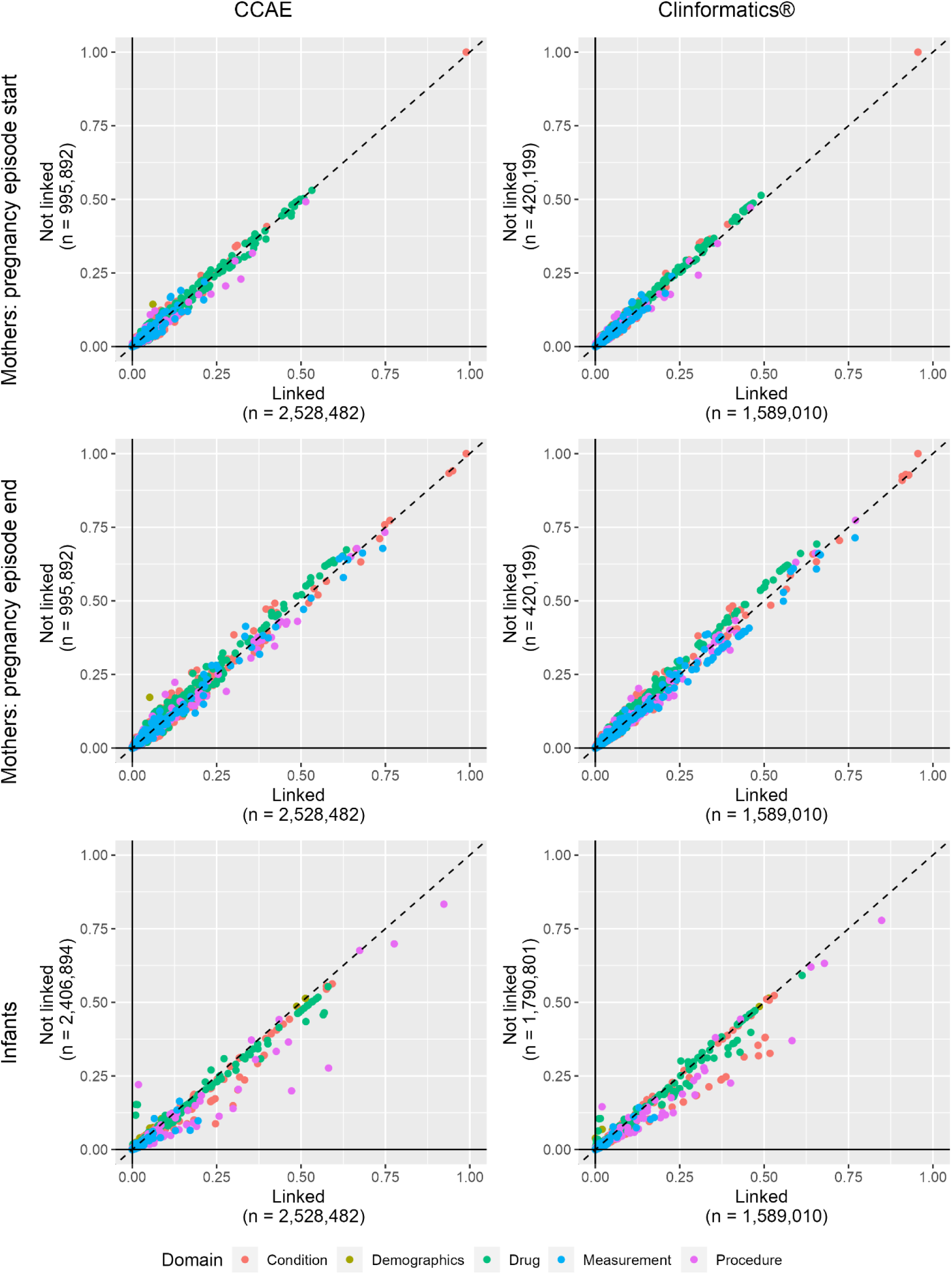
Demographic, drug exposure, condition, procedure, measurement, and visit occurrence covariate prevalence comparing linked vs. non-linked mother cohorts indexed at pregnancy episode start (row 1), linked vs non-linked mother cohorts indexed at pregnancy episode end (row 2) in IBM® Marketscan® Commercial Database and Optum’s de-identified Clinformatics® Data Mart Database. Primary algorithm implementation: all births, ±60-day pregnancy episode end/infant date-of-birth correspondence. CCAE: IBM® Marketscan® Commercial Database; Optum: Optum’s de-identified Clinformatics® Data Mart Database. The x-axes display the prevalence of each covariate in the linked populations and the y-axes display the prevalence of each covariate in the non-linked population. Data points that lay on the diagonal represent covariates that are equally prevalent in the linked and non-linked populations. Data points to the right the diagonal represent covariates that are more prevalent in the linked populations and those to the left are more prevalent in the non-linked populations.

The plots illustrate that the characteristics of linked and non-linked mothers were generally similar. However, infant characteristics, including conditions, measurements, drugs, and procedures were more prevalent among linked vs. non-linked infants. Large SMD covariates with greater prevalence among the linked infants included procedural billing records related to infant care, infant screening procedures, immunizations, and some conditions (see interactive web application to review all characteristics). We also observed a greater prevalence of birth-related covariates among linked infants than non-linked infants (e.g., “Single live birth”, ”Finding related to pregnancy”). Despite these differences, we still observed absolute SMDs of <0.1 for >99% of covariates across all algorithm implementations of each linked vs non-linked comparison in both databases where the number of covariate comparisons ranged from 58,611 (CCAE infants) to 68,368 (Clinformatics® mothers pregnancy end).

The final person and record counts for each of the 9 cohorts constructed by the 3 linkage algorithm implementations in each database are reported in Appendix Table C1. Result sets for the two algorithm sensitivity implementations are reported in Appendix Sections D and E. We observed similar stepwise attrition proportions across sensitivity implementations. Attrition proportions in the first births sensitivity implementation were greater in Step 3 because this is where first birth restrictions were made. There were no appreciable differences in linked vs. non-linked mother and infant characteristics across algorithm sensitivity implementations.

## Comment

### Principal Findings

We developed and implemented an algorithm to infer mother-infant links in two large US commercial healthcare databases that exhibited high linkage coverage and similar characteristics across linked vs. non-linked persons. This signifies generalizability of linked mother-infant pairs to commercially insured populations, which facilitates large-scale research on prenatal exposures and infant outcomes.

Our algorithm identified >3.4 million linked mothers and >4.1 million linked infants, which to our knowledge, makes these among the largest mother-infant linked cohorts constructed. Access to large, linked populations makes feasible the study of a wide range of prescription drug exposures, maternal and neonatal outcomes, and subgroups that are often unavailable in smaller linked populations[36, 37] and registries[38–40]. This approach requires fewer study resources compared to studies that require primary data collection[41].

### Results in the Context of What is Known

Across databases, linked mothers comprised 73.6% of all mothers with live births. In Clinformatics®, 77.3% of mothers were successfully linked to infants, which is lower but comparable to the 84% reported in a recent study using data from the same source with fewer linkage restrictions [18]. Despite similar methods, other linkage studies have reported mixed linkage coverage, suggesting that differences are due to data accuracy and/or availability variation across sources. Palmsten et al. linked Medicaid-enrolled mothers and infants and reported linkage coverage of 55.6% for inpatient deliveries, although with considerable variation by state (0-96%)[22], which the authors attribute to varying family identifier quality. A study in TRICARE enrollees in the Military Health System reported 90% of pregnancies ending in live births were linked with infants[42], which may be attributable to lower insurance coverage churn.

In our study, linked infants comprised 49.1% of all infants defined as persons 0 years of age at their observation period start. Contextualizing our linked infant coverage is difficult because most studies only report the proportion of linked pregnancies[18, 22]. However, Garbe et al. conducted a study using the German Pharmacoepidemiological Research Database (GePaRD), a claims database from four statutory health insurance providers, and reported that 77.3% of newborns were linked with mothers[43]. Additionally, a study among Medicaid enrollees in Tennessee reported 97% of infants were linked with a delivery, however such high coverage is likely explained by the use of vital record data with identifying information[37].

While our primary analysis used a ±60-day window between infant DOB and mothers’ pregnancy episode end to identify candidate links, in sensitivity analyses, we observed high correspondence at 7, 14, and 30 days, including same-day correspondence of 31.3% in CCAE and 67.4% in Clinformatics®. Overall correspondence was greater in Clinformatics®, which may be due to more accurate and specific DOB information. Increasing the correspondence window to 90 days increased the proportion of linked infants by 1.5% in CCAE and 0.2% in Clinformatics®, which we do not interpret as material because most of the correspondence occurs within ±30 days.

### Clinical and Research Implications

Characteristic comparisons between linked and non-linked mothers revealed similar demographic, health, and healthcare utilization profiles. Our linkage evaluation largely supports the generalizability of the linked mother population, having compared thousands of covariates between linked and non-linked mother cohorts and observing few differences. We found that SMDs were <0.1 for nearly all observed characteristics, suggesting that in a prenatal exposure study on a small, exposed subset of the linked mother population, systematic differences between the study sample and non-linked mothers to whom the results will apply will be minimal.

Despite substantial similarity between linked and non-linked infants, we observed more differences than when comparing mothers. Of note, linked infants had greater total healthcare utilization and prevalence of individual clinical events, including birth and infant-care related claims. Because our algorithm linked mothers to infants by a shared insurance ID within a defined temporal interval, candidate infants whose inferred DOB fell outside of that interval would be non-linked and less likely to have their clinical events captured in the database. This suggests that some billing records may be attributed to family members on other insurance plans among the non-linked populations. Still, this evaluation supports the generalizability of the linked infant population.

### Strengths and Limitations

A strength of our study is the rigorous linkage approach utilizing insurance ID in addition to delivery and birth procedure dates in large US claims databases representing the commercially insured population. Other studies have used related methods successfully[18, 22, 23, 43]. Further, we provide open-source code and a web-application for interactive characterization results, which provides valuable context for the external validity of future studies among linked populations. Additionally, our linked populations are larger than other mother-infant linked databases such as such as MEPREP (1,221,156 linked infants)[24, 25], the US Military Health System (827,753 linked live births)[23], GePaRD (250,355 linked live births), Medicaid Analytic Extract (1,173,280 linked pregnancies)[22], and Optum’s de-identified Clinformatics® Data Mart Database (524,227 linked live births)[18]. Lastly, developing a reproducible mother-child linkage algorithm in large administrative databases facilitates evidence generation in pregnant populations with improved rigor by avoiding recall, referral, and self-selection biases inherent to registry or other primary data collection studies of prenatal medication use[39].

Using administrative healthcare claims databases in pharmacoepidemiologic research has limitations. Erroneously coded or missing diagnostic, procedure, and drug dispensing records results in misclassification which may under- or over-estimate exposures, covariates, health outcomes, other clinical events, and healthcare utilization[44]. Subsequent information bias that can result from misclassification is underappreciated[45] and could bias findings of future drug safety studies. Still, developing reliable mother-infant linkages in large healthcare databases has increased the capacity to examine associations between rare prenatal drug exposures and infant outcomes with sufficient power. For example, prenatal use of antidepressants, stimulants, antihypertensive medications, and sulfonamides have been studied in relation to validated congenital anomalies[46–50]. This has yielded needed real-world evidence on the safety of prenatal exposures.

While we found few differences between linked and non-linked populations suggestive of high internal validity to the underlying commercially insured US population, our results do not necessarily ensure external validity to those covered under other types of insurance or lacking coverage. The data in this study are representative of people with US-based, employer-sponsored health insurance. Further, administrative healthcare databases include detailed outpatient drug dispensing records but provide fewer details on inpatient dispensing records, prescriptions, or administrations typically available in electronic medical records. Lastly, our study has not been validated. However validation of a similar algorithm developed in claims data among Medicaid beneficiaries showed high positive predictive value[51].

## Conclusions

Our study reinforces the shift towards implementing pharmacoepidemiology studies on prenatal drug exposures utilizing large electronic healthcare data as a supplement to traditional pregnancy registries. Our algorithm and evaluation demonstrate the ability to assemble large mother-infant linked cohorts for investigating prenatal drug exposure effects on infant outcomes.

## Data Availability

https://github.com/ohdsi-studies/MotherInfantLinkEval

## A. Algorithm sensitivity implementation descriptions

The primary linkage algorithm implementation identified all births for a given mother and required ±60-day correspondence between the infant’s date of birth (DOB) and the mother’s pregnancy episode end date as described in Step 3. The first sensitivity analysis limited results to first births per mother only and maintained the ±60-day correspondence between the infant’s DOB and the mother’s pregnancy episode end. The second sensitivity analysis identified all births for a given mother and increased the correspondence between the infant’s DOB and the mother’s pregnancy episode end to ±90-days. The intent of limiting to first births was to assess whether linked and non-linked mother characteristic differences depended on multiparous status. The intent of extending the pregnancy end/DOB correspondence was to increase linkage sensitivity given that some day of birth values were inferred as 1 from typical insurance enrollment start dates and new children may not be enrolled with coverage immediately after birth.

## B. Covariate construction

Mothers were characterized by age in years, index year (i.e., year in which pregnancy started/ended), index month, pre- and post-index database observation time in days, pregnancy episode length in days, and the Charlson Comorbidity Index (CCI)^1^. Infants were characterized by sex, index year, index month, and post-index observation time in days. Both mothers and infants were characterized by verbatim condition (SNOMED-CT) and aggregated condition group (SNOMED-CT clinical finding level) occurrences, drug ingredient level (Unified Medical Language System - RxNorm) and aggregated drug group (World Health Organization Anatomical Therapeutic Chemical Classification System 3^rd^, 4^th^, and 5^th^ levels) exposures, procedure occurrences, measurement occurrence (but not values), and counts of distinct conditions, drug ingredient exposures, procedures, and measurements. Health care utilization was characterized by proxy using visit counts and counts of specific visit types (outpatient, inpatient, emergency department). Continuous measures were reported with mean, standard deviation, and median and clinical event occurrences were reported as proportions. We chose pregnancy episode end as the index date for linked infants rather than the infant’s inferred DOB because live birth procedures are well-captured in claims data with reliable dates, whereas DOB information is limited or redacted as part of enrollment and de-identification and enrollment date is an imprecise birth marker.

## C. Person and record counts

1) Linked mothers (pregnancy episode start)

Mothers linked to ≥1 infant indexed at pregnancy episode start

2) Linked mothers (pregnancy episode end)

Mothers linked to ≥1 infant indexed at pregnancy episode end

3) Linked infants

Infants linked to a mother indexed at pregnancy episode end

4) Not linked mothers (pregnancy episode start)

Mothers not linked to an infant indexed at pregnancy episode start

5) Not linked mothers (pregnancy episode end)

Mothers not linked to an infant indexed at pregnancy episode end

6) Not linked infants

Infants not linked to a mother indexed at inferred DOB

7) All candidate mothers (pregnancy episode start)

Mothers without infant linkage restrictions indexed at pregnancy episode start

8) All candidate mothers (pregnancy episode end)

Mothers without infant linkage restrictions indexed at pregnancy episode end

9) All candidate infants

Infants without linkage restrictions indexed at DOB

**Table C1.**
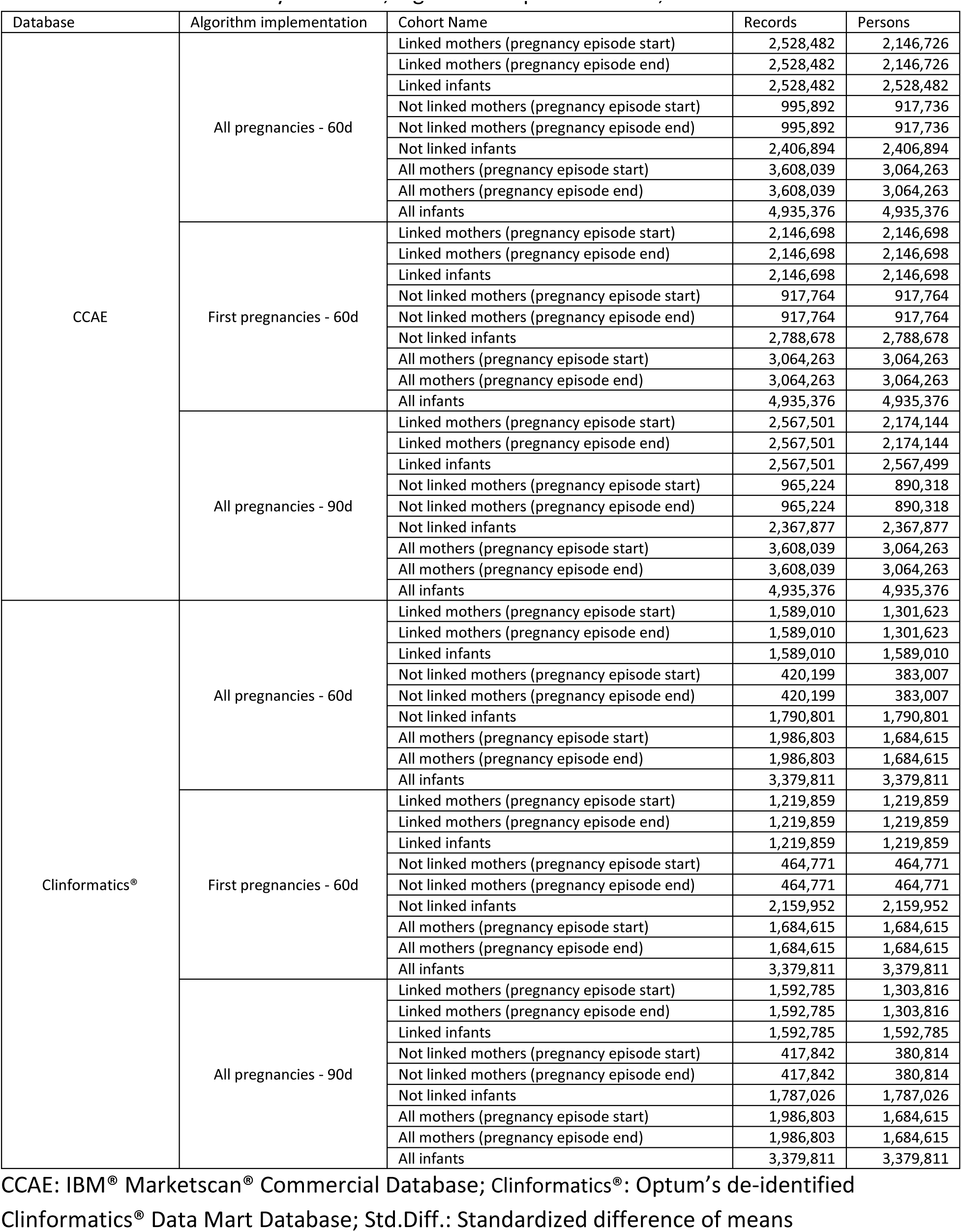
Person and record counts by database, algorithm implementation, and cohort

| Database | Algorithm implementation | Cohort Name | Records | Persons |
| --- | --- | --- | --- | --- |
| CCAE | All pregnancies - 60d | Linked mothers (pregnancy episode start) | 2,528,482 | 2,146,726 |
|  |  | Linked mothers (pregnancy episode end) | 2,528,482 | 2,146,726 |
|  |  | Linked infants | 2,528,482 | 2,528,482 |
|  |  | Not linked mothers (pregnancy episode start) | 995,892 | 917,736 |
|  |  | Not linked mothers (pregnancy episode end) | 995,892 | 917,736 |
|  |  | Not linked infants | 2,406,894 | 2,406,894 |
|  |  | All mothers (pregnancy episode start) | 3,608,039 | 3,064,263 |
|  |  | All mothers (pregnancy episode end) | 3,608,039 | 3,064,263 |
|  |  | All infants | 4,935,376 | 4,935,376 |
|  | First pregnancies - 60d | Linked mothers (pregnancy episode start) | 2,146,698 | 2,146,698 |
|  |  | Linked mothers (pregnancy episode end) | 2,146,698 | 2,146,698 |
|  |  | Linked infants | 2,146,698 | 2,146,698 |
|  |  | Not linked mothers (pregnancy episode start) | 917,764 | 917,764 |
|  |  | Not linked mothers (pregnancy episode end) | 917,764 | 917,764 |
|  |  | Not linked infants | 2,788,678 | 2,788,678 |
|  |  | All mothers (pregnancy episode start) | 3,064,263 | 3,064,263 |
|  |  | All mothers (pregnancy episode end) | 3,064,263 | 3,064,263 |
|  |  | All infants | 4,935,376 | 4,935,376 |
|  | All pregnancies - 90d | Linked mothers (pregnancy episode start) | 2,567,501 | 2,174,144 |
|  |  | Linked mothers (pregnancy episode end) | 2,567,501 | 2,174,144 |
|  |  | Linked infants | 2,567,501 | 2,567,499 |
|  |  | Not linked mothers (pregnancy episode start) | 965,224 | 890,318 |
|  |  | Not linked mothers (pregnancy episode end) | 965,224 | 890,318 |
|  |  | Not linked infants | 2,367,877 | 2,367,877 |
|  |  | All mothers (pregnancy episode start) | 3,608,039 | 3,064,263 |
|  |  | All mothers (pregnancy episode end) | 3,608,039 | 3,064,263 |
|  |  | All infants | 4,935,376 | 4,935,376 |
| Clinformatics® | All pregnancies - 60d | Linked mothers (pregnancy episode start) | 1,589,010 | 1,301,623 |
|  |  | Linked mothers (pregnancy episode end) | 1,589,010 | 1,301,623 |
|  |  | Linked infants | 1,589,010 | 1,589,010 |
|  |  | Not linked mothers (pregnancy episode start) | 420,199 | 383,007 |
|  |  | Not linked mothers (pregnancy episode end) | 420,199 | 383,007 |
|  |  | Not linked infants | 1,790,801 | 1,790,801 |
|  |  | All mothers (pregnancy episode start) | 1,986,803 | 1,684,615 |
|  |  | All mothers (pregnancy episode end) | 1,986,803 | 1,684,615 |
|  |  | All infants | 3,379,811 | 3,379,811 |
|  | First pregnancies - 60d | Linked mothers (pregnancy episode start) | 1,219,859 | 1,219,859 |
|  |  | Linked mothers (pregnancy episode end) | 1,219,859 | 1,219,859 |
|  |  | Linked infants | 1,219,859 | 1,219,859 |
|  |  | Not linked mothers (pregnancy episode start) | 464,771 | 464,771 |
|  |  | Not linked mothers (pregnancy episode end) | 464,771 | 464,771 |
|  |  | Not linked infants | 2,159,952 | 2,159,952 |
|  |  | All mothers (pregnancy episode start) | 1,684,615 | 1,684,615 |
|  |  | All mothers (pregnancy episode end) | 1,684,615 | 1,684,615 |
|  |  | All infants | 3,379,811 | 3,379,811 |
|  | All pregnancies - 90d | Linked mothers (pregnancy episode start) | 1,592,785 | 1,303,816 |
|  |  | Linked mothers (pregnancy episode end) | 1,592,785 | 1,303,816 |
|  |  | Linked infants | 1,592,785 | 1,592,785 |
|  |  | Not linked mothers (pregnancy episode start) | 417,842 | 380,814 |
|  |  | Not linked mothers (pregnancy episode end) | 417,842 | 380,814 |
|  |  | Not linked infants | 1,787,026 | 1,787,026 |
|  |  | All mothers (pregnancy episode start) | 1,986,803 | 1,684,615 |
|  |  | All mothers (pregnancy episode end) | 1,986,803 | 1,684,615 |
|  |  | All infants | 3,379,811 | 3,379,811 |

CCAE: IBM® MarketScan® Commercial Database; Clinformatics®: Optum's de-identified Clinformatics® Data Mart Database; Std.Diff.: Standardized difference of means

## D. Linkage algorithm sensitivity implementation 1 evaluation results

**Figure D1.**
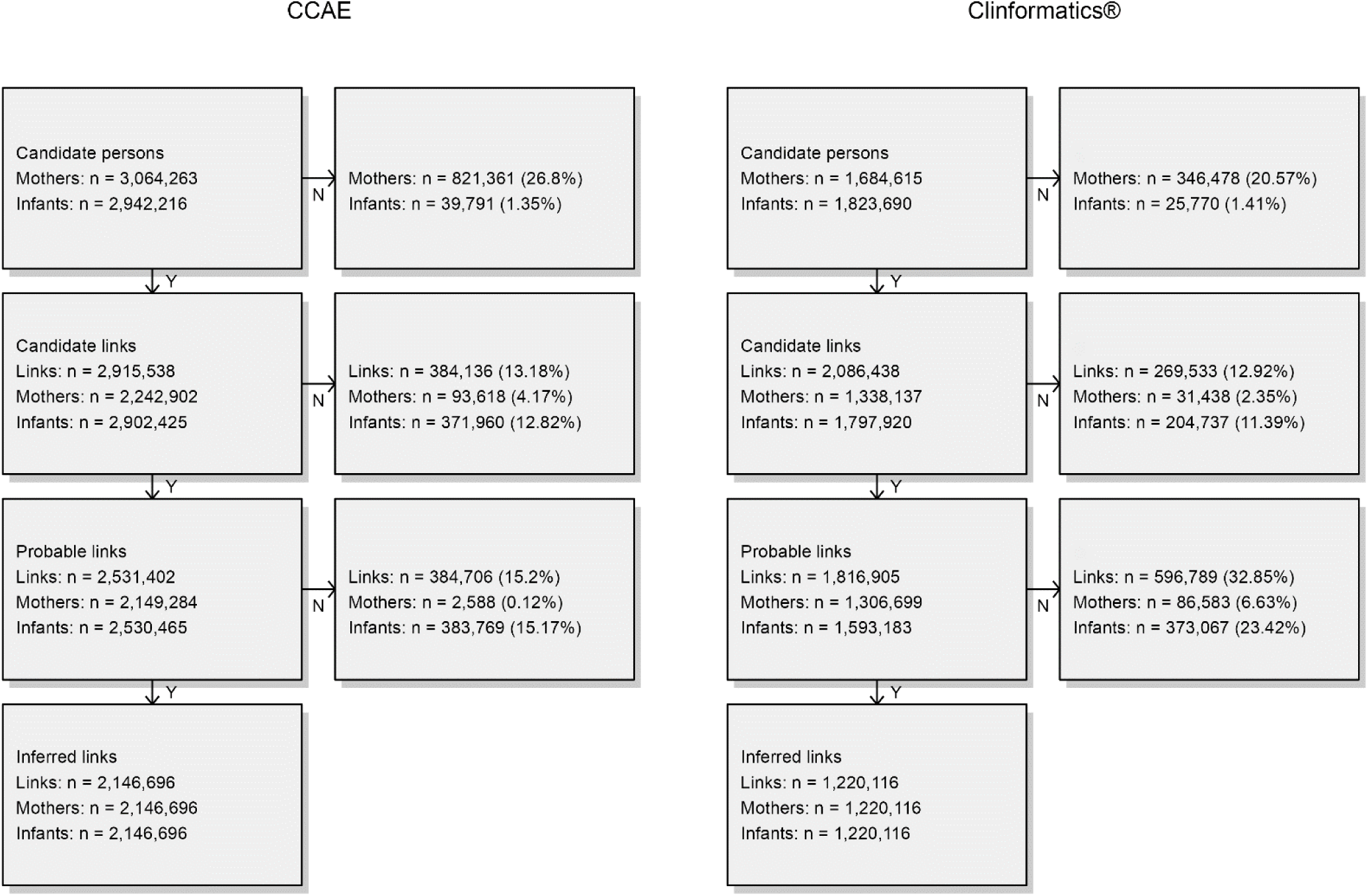
Mother-infant linkage algorithm attrition diagram. Sensitivity algorithm implementation 1: first births, ±60-day pregnancy episode end/infant date-of-birth correspondence. CCAE: IBM® Marketscan® Commercial Database; Clinformatics®: Optum’s de-identified Clinformatics® Data Mart Database

**Figure D2.**
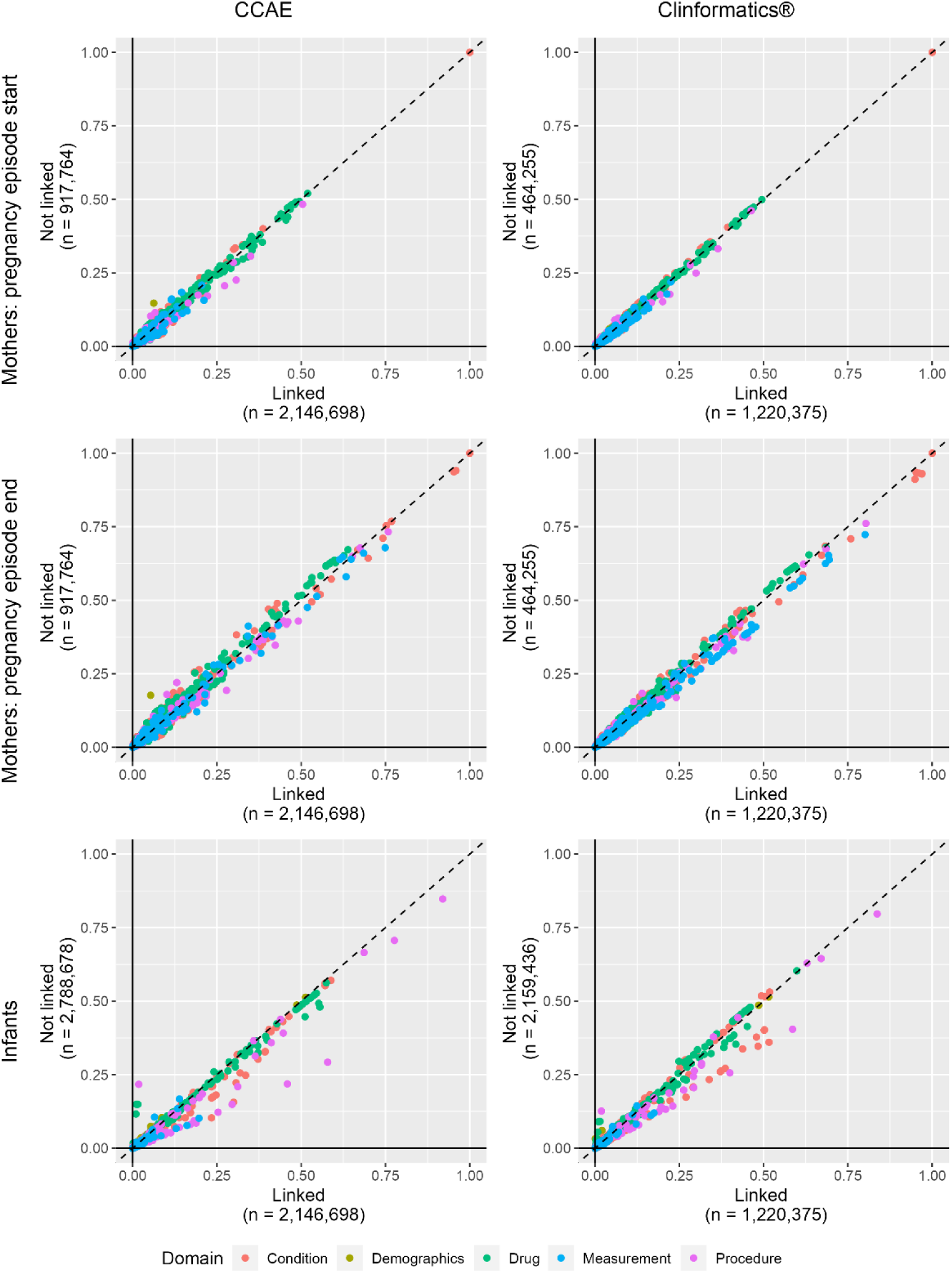
Demographic, drug exposure, condition, procedure, measurement, and visit occurrence covariate prevalence comparing linked vs. non-linked mother cohorts indexed at pregnancy episode start (row 1), linked vs non-linked mother cohorts indexed at pregnancy episode end (row 2). Sensitivity algorithm implementation 1: first births, ±60-day pregnancy episode end/infant date-of-birth correspondence. CCAE: IBM® Marketscan® Commercial Database; Clinformatics®: Optum’s de-identified Clinformatics® Data Mart Database.

**Table D1.** Selected characteristics and standardized differences of linked and non-linked mothers in IBM® Marketscan® Commercial Database and Optum’s de-identified Clinformatics® Data Mart Database. Mother characteristics were measured on the pregnancy episode start date (index year and month, age) or during the 365-day period before and including the pregnancy episode start date (distinct event occurrence counts). Sensitivity algorithm implementation 1: first births, ±60-day pregnancy episode end/infant date-of-birth correspondence.

| Characteristic | CCAЕ |  |  | Clinformatics® |  |  |
| --- | --- | --- | --- | --- | --- | --- |
|  | Linked<br>% (n = 2,146,698) | Non-linked<br>% (n = 917,764) | Std.Diff | Linked<br>% (n = 1,219,859) | Non-linked<br>% (n = 464,771) | Std.Diff |
| Index year |  |  |  |  |  |  |
| 2000 | 0.72 | 0.78 | 0.007 | 1.35 | 2.23 | 0.066 |
| 2001 | 1.08 | 1.21 | 0.013 | 4.01 | 5.53 | 0.071 |
| 2002 | 2.15 | 2.16 | 0 | 4.84 | 6.42 | 0.069 |
| 2003 | 3.46 | 2.77 | -0.04 | 4.92 | 5.79 | 0.038 |
| 2004 | 4.34 | 3.53 | -0.042 | 4.88 | 4.96 | 0.003 |
| 2005 | 4.16 | 3.37 | -0.042 | 6.18 | 4.73 | -0.064 |
| 2006 | 4.92 | 4.24 | -0.033 | 6.31 | 5.28 | -0.044 |
| 2007 | 5 | 4.47 | -0.025 | 6.31 | 5.08 | -0.053 |
| 2008 | 6.21 | 5.39 | -0.035 | 5.85 | 4.75 | -0.049 |
| 2009 | 6.28 | 5.98 | -0.012 | 5.01 | 5.09 | 0.004 |
| 2010 | 7.6 | 6.62 | -0.038 | 4.4 | 5.65 | 0.057 |
| 2011 | 8.24 | 8.49 | 0.009 | 4.21 | 5.94 | 0.079 |
| 2012 | 6.7 | 7.7 | 0.038 | 4.68 | 4.85 | 0.008 |
| 2013 | 6.78 | 7.18 | 0.016 | 4.59 | 4.32 | -0.013 |
| 2014 | 5.68 | 6.67 | 0.041 | 4.32 | 4.19 | -0.007 |
| 2015 | 5.05 | 5.77 | 0.032 | 4.56 | 4.36 | -0.01 |
| 2016 | 4.66 | 5.21 | 0.025 | 4.69 | 4.43 | -0.012 |
| 2017 | 4.62 | 4.9 | 0.013 | 4.85 | 4.47 | -0.018 |
| 2018 | 4.91 | 5.08 | 0.008 | 4.89 | 4.22 | -0.032 |
| 2019 | 4.24 | 4.59 | 0.017 | 4.57 | 3.96 | -0.03 |
| 2020 | 3.19 | 3.9 | 0.038 | 4.45 | 3.63 | -0.041 |
| 2021 | 0 | 0 | 0.007 | 0.12 | 0.14 | 0.007 |
| Index month |  |  |  |  |  |  |
| 1 | 10.2 | 9.8 | -0.013 | 9.1 | 9.19 | 0.003 |
| 2 | 7.73 | 10.88 | 0.109 | 7.87 | 8.26 | 0.014 |
| 3 | 6.26 | 14.69 | 0.278 | 7.94 | 8.96 | 0.037 |
| 4 | 7.49 | 8.42 | 0.034 | 7.45 | 7.63 | 0.007 |
| 5 | 8.5 | 7.01 | -0.056 | 8.27 | 8.04 | -0.008 |
| 6 | 8.36 | 6.72 | -0.062 | 8.05 | 7.87 | -0.007 |
| 7 | 8.8 | 6.81 | -0.074 | 8.38 | 8.15 | -0.008 |
| 8 | 8.69 | 6.76 | -0.072 | 8.3 | 8.05 | -0.009 |
| 9 | 8.6 | 7.02 | -0.059 | 8.47 | 8.26 | -0.008 |
| 10 | 8.65 | 7.35 | -0.048 | 8.77 | 8.5 | -0.01 |
| 11 | 8.41 | 7.16 | -0.047 | 8.66 | 8.4 | -0.009 |
| 12 | 8.32 | 7.39 | -0.035 | 8.72 | 8.67 | -0.002 |
| Age (years) |  |  |  |  |  |  |
| Mean | 31.01 | 27.38 | -0.449 | 30.64 | 28.37 | -0.274 |
| Std. deviation | 4.74 | 6.52 |  | 4.95 | 6.63 |  |
| Median | 31 | 27 |  | 31 | 28 |  |
| Prior observation time (days) |  |  |  |  |  |  |
| Mean | 590.75 | 750.23 | 0.146 | 581.03 | 655.96 | 0.075 |
| Std. deviation | 607.94 | 904.84 |  | 619.27 | 779.33 |  |
| Median | 402 | 441 |  | 391 | 403 |  |
| Post observation time (days) |  |  |  |  |  |  |
| Mean | 1335.44 | 955.32 | -0.254 | 1172.59 | 1021.04 | -0.106 |
| Std. deviation | 1190.85 | 906.25 |  | 1062.44 | 950.33 |  |
| Median | 908 | 643 |  | 784 | 686 |  |
| Pregnancy episode length (days) |  |  |  |  |  |  |
| Mean | 273.37 | 270.19 | -0.107 | 273.1 | 271.46 | -0.056 |
| Std. deviation | 18.5 | 23.19 |  | 18.77 | 22.26 |  |
| Median | 279 | 277 |  | 278 | 279 |  |
| Distinct conditions |  |  |  |  |  |  |
| Mean | 6.3 | 6.33 | 0.003 | 6.76 | 6.83 | 0.006 |
| Std. deviation | 6.37 | 6.78 |  | 6.99 | 7.39 |  |
| Median | 4 | 4 |  | 5 | 4 |  |
| Distinct drug ingredients |  |  |  |  |  |  |
| Mean | 4.57 | 4.48 | -0.012 | 4.18 | 4.18 | 0.001 |
| Std. deviation | 5.41 | 5.43 |  | 5.18 | 5.31 |  |
| Median | 3 | 3 |  | 3 | 3 |  |
| Distinct procedures |  |  |  |  |  |  |
| Mean | 7.13 | 6.3 | -0.082 | 6.98 | 6.6 | -0.036 |
| Std. deviation | 7.35 | 7.06 |  | 7.53 | 7.46 |  |
| Median | 5 | 4 |  | 5 | 5 |  |
| Distinct measurements |  |  |  |  |  |  |
| Mean | 6.57 | 6.01 | -0.033 | 13.79 | 11.6 | -0.059 |
| Std. deviation | 12.03 | 11.75 |  | 27.65 | 25.14 |  |
| Median | 3 | 2 |  | 3 | 3 |  |
| Distinct visit types |  |  |  |  |  |  |
| Outpatient Visit |  |  |  |  |  |  |
| Mean | 6.28 | 5.3 | -0.079 | 5.98 | 5.59 | -0.031 |
| Std. deviation | 9.35 | 8.25 |  | 9.09 | 8.71 |  |
| Median | 3 | 3 |  | 3 | 3 |  |
| Inpatient Visit |  |  |  |  |  |  |
| Mean | 0.08 | 0.08 | 0.001 | 0.07 | 0.08 | 0.005 |
| Std. deviation | 1.22 | 1.44 |  | 0.72 | 1.02 |  |
| Median | 0 | 0 |  | 0 | 0 |  |
| Emergency Room Visit |  |  |  |  |  |  |
| Mean | 0.16 | 0.31 | 0.083 | 0.36 | 0.31 | -0.007 |
| Std. deviation | 1.13 | 1.48 |  | 5.62 | 4.97 |  |
| Median | 0 | 0 |  | 0 | 0 |  |
| Charlson comorbidity index |  |  |  |  |  |  |
| Mean | 0.17 | 0.18 | 0.012 | 0.2 | 0.21 | 0.012 |
| Std. deviation | 0.83 | 0.88 |  | 0.91 | 0.99 |  |
| Median | 0 | 0 |  | 0 | 0 |  |

CCAE: IBM® MarketScan® Commercial Database; Clinformatics®: Optum's de-identified Clinformatics® Data Mart Database; Std.Diff.: Standardized difference of means

**Table D2.** Selected characteristics and standardized differences of linked and non-linked mothers in IBM® Marketscan® Commercial Database and Optum’s de-identified Clinformatics® Data Mart Database. Mother characteristics were measured on the pregnancy episode end date (index year and month, age) or during the 365-day period before and including the pregnancy episode end date (distinct event occurrence counts). Sensitivity algorithm implementation 1: first births, ±60-day pregnancy episode end/infant date-of-birth correspondence.

| Characteristic | CCAE |  |  | Clinformatics® |  |  |
| --- | --- | --- | --- | --- | --- | --- |
|  | Linked<br>% (n = 2,146,698) | Non-linked<br>% (n = 917,764) | Std.Diff | Linked<br>% (n = 1,220,375) | Non-linked<br>% (n = 464,255) | Std.Diff |
| Index year |  |  |  |  |  |  |
| 2000 | 0.11 | 0.33 | 0.047 | 0.01 | 0.02 | 0.012 |
| 2001 | 0.75 | 0.93 | 0.02 | 2.03 | 3.35 | 0.082 |
| 2002 | 1.31 | 1.61 | 0.025 | 4.47 | 6.15 | 0.075 |
| 2003 | 2.49 | 2.39 | -0.006 | 4.93 | 6.43 | 0.065 |
| 2004 | 3.67 | 3 | -0.037 | 4.88 | 5.51 | 0.029 |
| 2005 | 4.38 | 3.57 | -0.041 | 5.12 | 4.53 | -0.028 |
| 2006 | 4.22 | 3.48 | -0.039 | 6.41 | 5.03 | -0.059 |
| 2007 | 4.97 | 4.33 | -0.03 | 6.31 | 5.29 | -0.044 |
| 2008 | 5.35 | 4.84 | -0.023 | 6.25 | 4.98 | -0.055 |
| 2009 | 6.53 | 5.54 | -0.042 | 5.69 | 4.67 | -0.046 |
| 2010 | 6.27 | 6.43 | 0.007 | 4.69 | 5.45 | 0.035 |
| 2011 | 7.8 | 6.95 | -0.033 | 4.32 | 5.74 | 0.065 |
| 2012 | 8.08 | 9.05 | 0.035 | 4.24 | 5.9 | 0.076 |
| 2013 | 6.52 | 6.91 | 0.016 | 4.89 | 4.45 | -0.021 |
| 2014 | 6.79 | 7.3 | 0.02 | 4.45 | 4.25 | -0.01 |
| 2015 | 5.21 | 5.97 | 0.033 | 4.41 | 4.25 | -0.008 |
| 2016 | 5.14 | 5.8 | 0.029 | 4.71 | 4.45 | -0.013 |
| 2017 | 4.66 | 5.02 | 0.017 | 4.7 | 4.45 | -0.012 |
| 2018 | 4.68 | 4.98 | 0.014 | 4.86 | 4.41 | -0.022 |
| 2019 | 4.77 | 4.93 | 0.007 | 4.87 | 4.18 | -0.033 |
| 2020 | 4.16 | 4.48 | 0.016 | 4.47 | 3.83 | -0.032 |
| 2021 | 2.14 | 2.15 | 0 | 3.29 | 2.67 | -0.037 |
| Index month |  |  |  |  |  |  |
| 1 | 8.17 | 6.66 | -0.058 | 7.59 | 7.47 | -0.005 |
| 2 | 7.79 | 6.14 | -0.065 | 7.44 | 7.1 | -0.013 |
| 3 | 8.68 | 6.76 | -0.072 | 8.31 | 8.1 | -0.008 |
| 4 | 8.48 | 6.56 | -0.073 | 8.11 | 7.89 | -0.008 |
| 5 | 8.86 | 6.81 | -0.076 | 8.51 | 8.18 | -0.012 |
| 6 | 8.58 | 7.12 | -0.054 | 8.43 | 8.23 | -0.007 |
| 7 | 8.64 | 7.39 | -0.046 | 8.76 | 8.49 | -0.009 |
| 8 | 8.66 | 7.39 | -0.047 | 8.93 | 8.75 | -0.006 |
| 9 | 8.44 | 7.55 | -0.033 | 8.67 | 8.58 | -0.003 |
| 10 | 9.81 | 9 | -0.028 | 8.79 | 8.91 | 0.004 |
| 11 | 8.56 | 10.94 | 0.081 | 8.35 | 8.62 | 0.01 |
| 12 | 5.32 | 17.68 | 0.395 | 8.13 | 9.69 | 0.055 |
| Age (years) |  |  |  |  |  |  |
| Mean | 31.77 | 27.99 | -0.471 | 31.38 | 29.09 | -0.278 |
| Std. deviation | 4.74 | 6.47 |  | 4.94 | 6.62 |  |
| Median | 32 | 28 |  | 31 | 29 |  |
| Prior observation time (days) |  |  |  |  |  |  |
| Mean | 864.12 | 1020.43 | 0.143 | 854.17 | 927.43 | 0.074 |
| Std. deviation | 607.63 | 904.84 |  | 619.29 | 778.87 |  |
| Median | 675 | 714 |  | 665 | 675 |  |
| Post observation time (days) |  |  |  |  |  |  |
| Mean | 1062.07 | 685.13 | -0.252 | 899.9 | 748.3 | -0.107 |
| Std. deviation | 1189.86 | 905.78 |  | 1061.97 | 947.24 |  |
| Median | 635 | 372 |  | 512 | 414 |  |
| Distinct conditions |  |  |  |  |  |  |
| Mean | 20.01 | 20.6 | 0.039 | 21.28 | 20.79 | -0.031 |
| Std. deviation | 10.2 | 11.26 |  | 10.75 | 11.35 |  |
| Median | 18 | 18 |  | 19 | 18 |  |
| Distinct drug ingredients |  |  |  |  |  |  |
| Mean | 5.35 | 5.61 | 0.034 | 5.67 | 5.7 | 0.004 |
| Std. deviation | 5.38 | 5.6 |  | 5.42 | 5.68 |  |
| Median | 4 | 4 |  | 4 | 4 |  |
| Distinct procedures |  |  |  |  |  |  |
| Mean | 17.72 | 16.8 | -0.076 | 19.52 | 18.5 | -0.081 |
| Std. deviation | 8.55 | 8.52 |  | 8.81 | 9.01 |  |
| Median | 16 | 15 |  | 18 | 17 |  |
| Distinct measurements |  |  |  |  |  |  |
| Mean | 21.24 | 21.01 | -0.01 | 46.41 | 40.93 | -0.108 |
| Std. deviation | 15.77 | 16.67 |  | 36.8 | 35.2 |  |
| Median | 18 | 18 |  | 36 | 27 |  |
| Distinct visit types |  |  |  |  |  |  |
| Outpatient Visit |  |  |  |  |  |  |
| Mean | 15.46 | 14.23 | -0.087 | 15.55 | 14.6 | -0.062 |
| Std. deviation | 10.52 | 9.52 |  | 10.89 | 10.7 |  |
| Median | 13 | 12 |  | 13 | 12 |  |
| Inpatient Visit |  |  |  |  |  |  |
| Mean | 1.13 | 1.13 | 0.002 | 1.13 | 1.12 | -0.016 |
| Std. deviation | 0.63 | 0.69 |  | 0.55 | 0.62 |  |
| Median | 1 | 1 |  | 1 | 1 |  |
| Emergency Room Visit |  |  |  |  |  |  |
| Mean | 0.34 | 0.65 | 0.139 | 1.03 | 0.9 | -0.019 |
| Std. deviation | 1.32 | 1.81 |  | 5.01 | 4.64 |  |
| Median | 0 | 0 |  | 0 | 0 |  |
| Charlson comorbidity index |  |  |  |  |  |  |
| Mean | 0.27 | 0.28 | 0.007 | 0.31 | 0.31 | 0.004 |
| Std. deviation | 0.89 | 0.94 |  | 0.97 | 1.05 |  |
| Median | 0 | 0 |  | 0 | 0 |  |

CCAE: IBM® MarketScan® Commercial Database; Clinformatics®: Optum's de-identified Clinformatics® Data Mart Database; Std.Diff.: Standardized difference of means

**Table D3.**
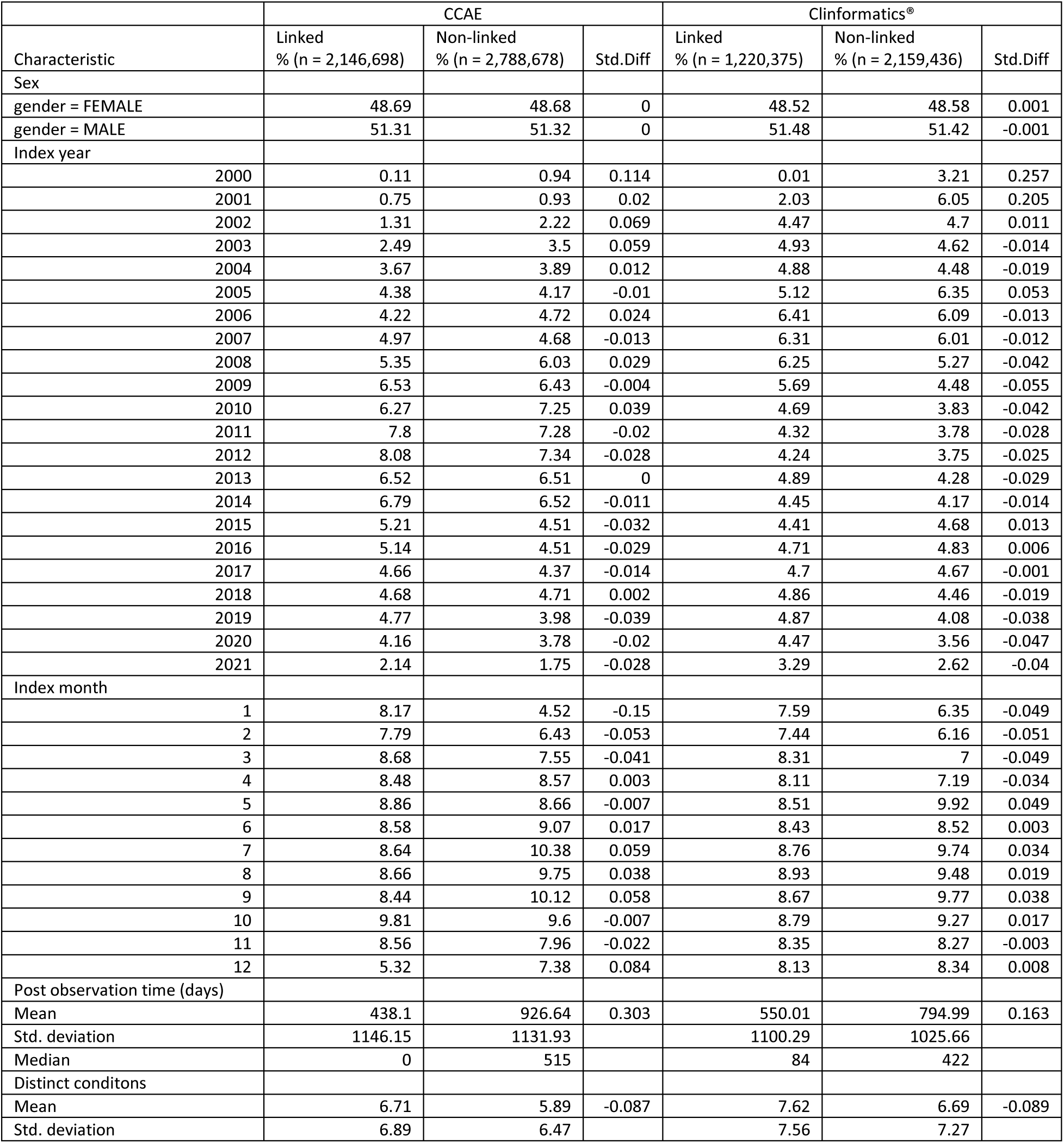
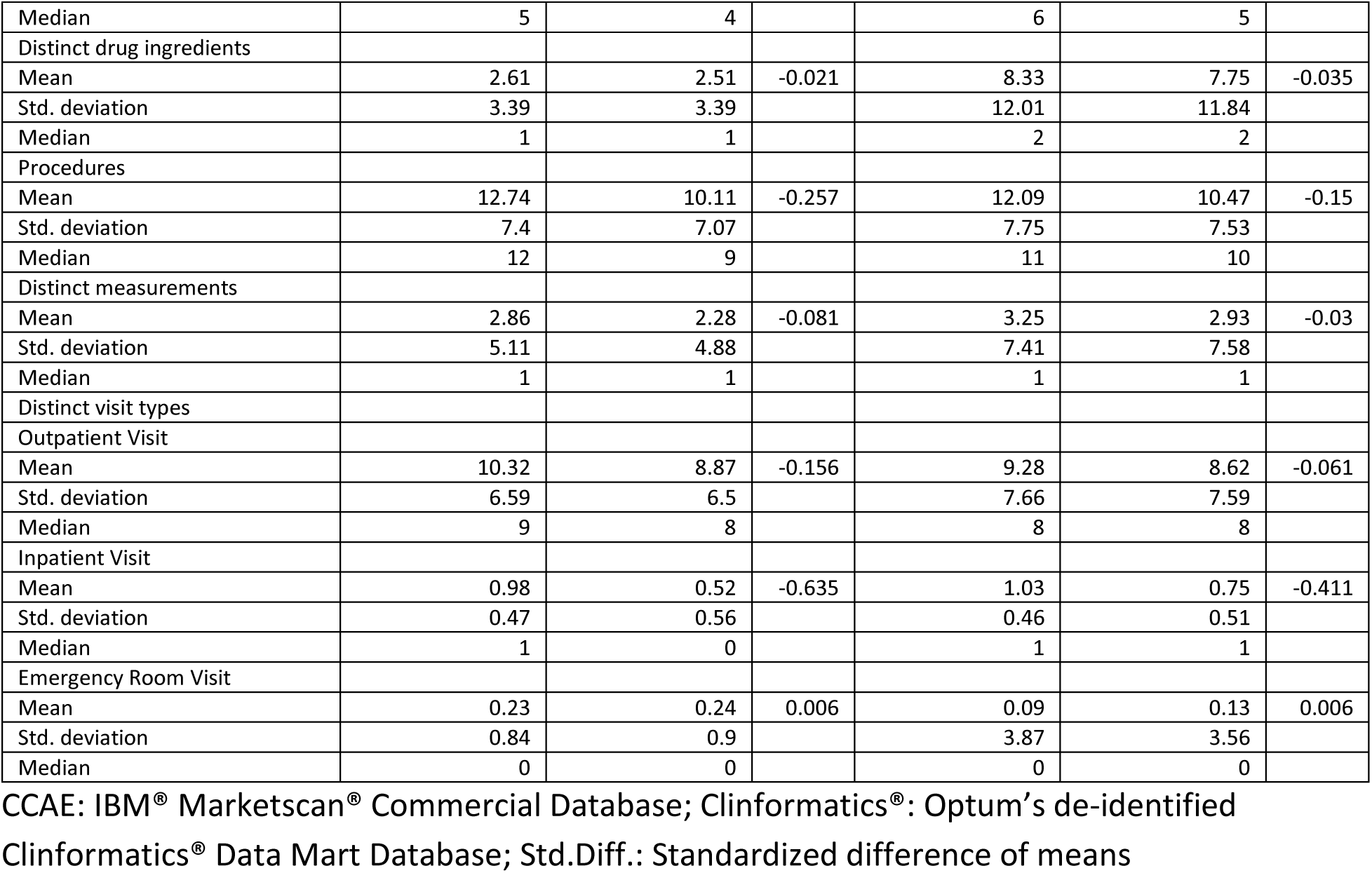
Selected characteristics and standardized differences of linked and non-linked infants in IBM® Marketscan® Commercial Database and Optum’s de-identified Clinformatics® Data Mart Database. Infant characteristics were measured on the pregnancy episode end date (index year and month, age, sex) or during the 365-day period after and including the pregnancy episode end date (distinct event occurrence counts). Sensitivity algorithm implementation 1: first births, ±60-day pregnancy episode end/infant date-of-birth correspondence.

## E. Linkage algorithm sensitivity implementation 2 evaluation results

**Figure E1.**
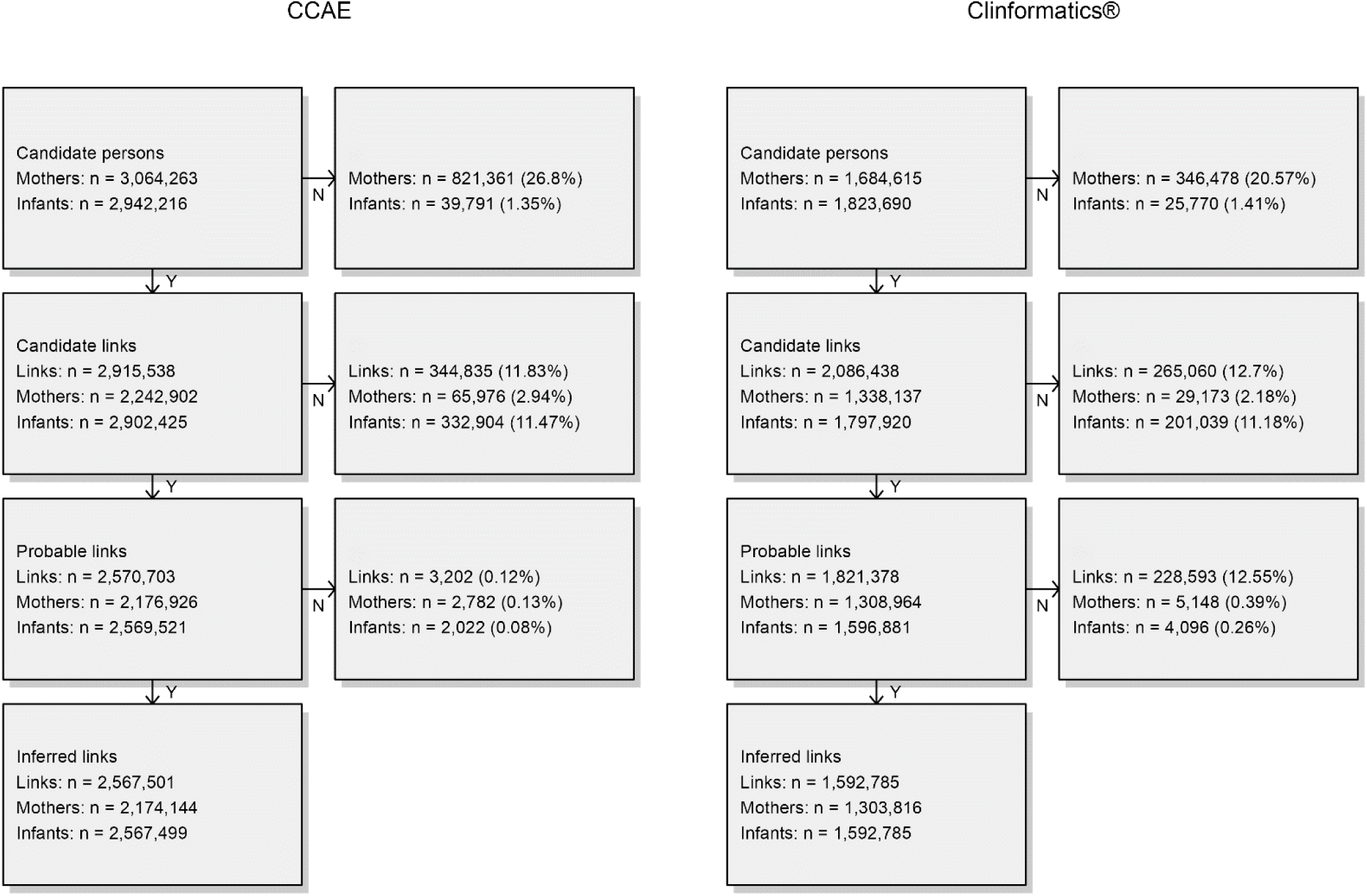
Mother-infant linkage algorithm attrition diagram. Sensitivity algorithm implementation 2: all births, ±90-day pregnancy episode end/infant date-of-birth correspondence. CCAE: IBM® Marketscan® Commercial Database; Clinformatics®: Optum’s de-identified Clinformatics® Data Mart Database

**Figure E2.**
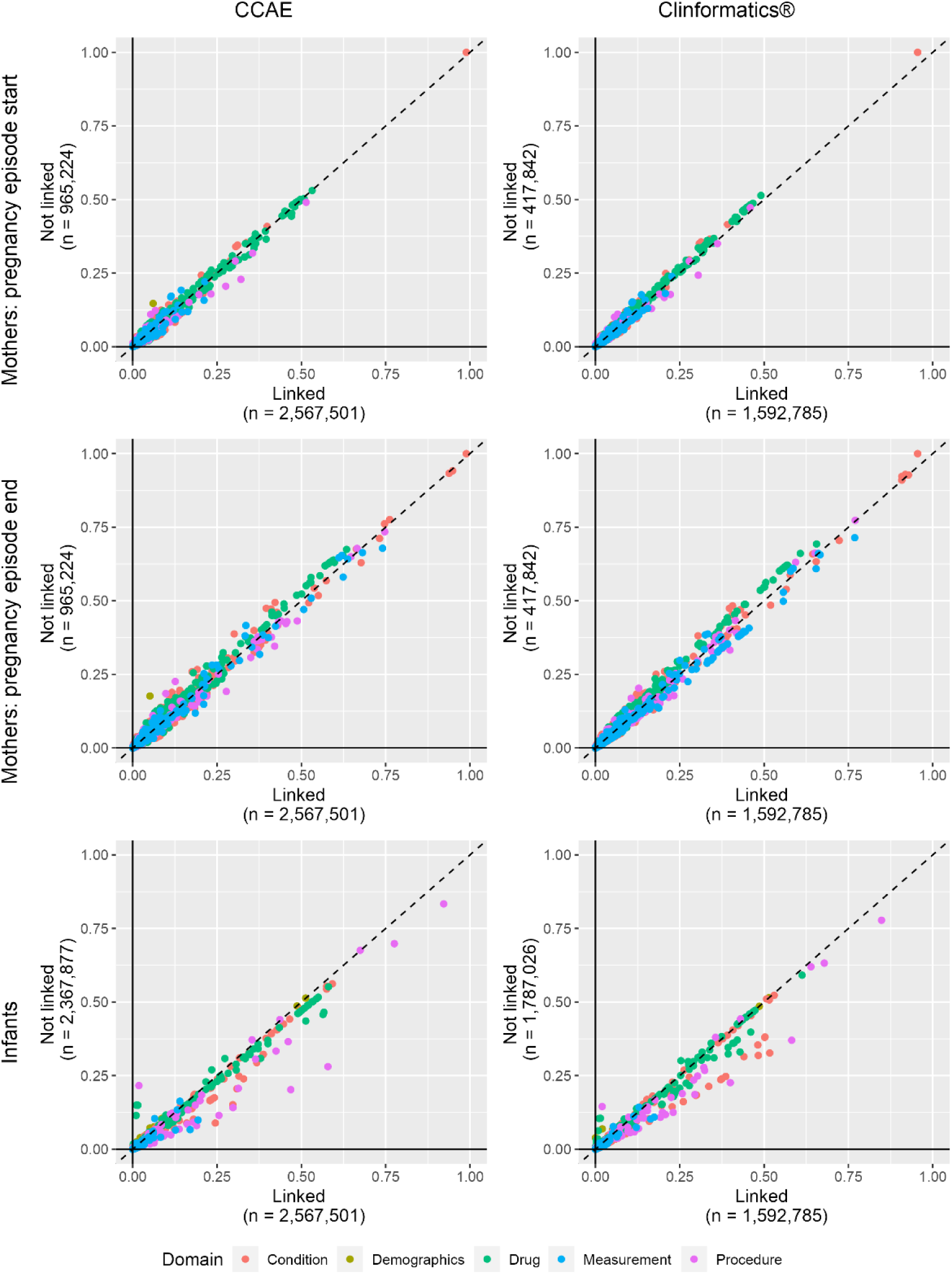
Demographic, drug exposure, condition, procedure, measurement, and visit occurrence covariate prevalence comparing linked vs. non-linked mother cohorts indexed at pregnancy episode start (row 1), linked vs non-linked mother cohorts indexed at pregnancy episode end (row 2). Sensitivity algorithm implementation 2: all births, ±90-day pregnancy episode end/infant date-of-birth correspondence. CCAE: IBM® Marketscan® Commercial Database; Clinformatics®: Optum’s de-identified Clinformatics® Data Mart Database.

**Table E1.** Selected characteristics and standardized differences of linked and non-linked mothers in IBM® Marketscan® Commercial Database and Optum’s de-identified Clinformatics® Data Mart Database. Mother characteristics were measured on the pregnancy episode start date (index year and month, age) or during the 365-day period before and including the pregnancy episode start date (distinct event occurrence counts). Sensitivity algorithm implementation 2: all births, ±90-day pregnancy episode end/infant date-of-birth correspondence.

| Characteristic | CCAЕ |  |  | Clinformatics® |  |  |
| --- | --- | --- | --- | --- | --- | --- |
|  | Linked<br>% (n = 2,567,501) | Non-linked<br>% (n = 965,224) | Std.Diff | Linked<br>% (n = 1,592,785) | Non-linked<br>% (n = 417,842) | Std.Diff |
| Index year |  |  |  |  |  |  |
| 2000 | 0.63 | 0.69 | 0.007 | 1.36 | 1.38 | 0.002 |
| 2001 | 0.97 | 1.05 | 0.008 | 3.84 | 3.76 | -0.004 |
| 2002 | 1.95 | 1.97 | 0.001 | 4.72 | 4.53 | -0.009 |
| 2003 | 3.15 | 2.54 | -0.037 | 4.84 | 4.64 | -0.009 |
| 2004 | 4.05 | 3.3 | -0.04 | 4.8 | 4.48 | -0.015 |
| 2005 | 3.99 | 3.19 | -0.043 | 6.18 | 4.08 | -0.095 |
| 2006 | 4.77 | 4.03 | -0.036 | 6.3 | 4.76 | -0.067 |
| 2007 | 4.95 | 4.3 | -0.031 | 6.31 | 4.82 | -0.065 |
| 2008 | 6.07 | 5.23 | -0.036 | 5.92 | 4.64 | -0.057 |
| 2009 | 6.18 | 5.82 | -0.015 | 5.12 | 5.18 | 0.003 |
| 2010 | 7.43 | 6.48 | -0.037 | 4.54 | 6 | 0.065 |
| 2011 | 8.13 | 8.42 | 0.011 | 4.38 | 6.46 | 0.092 |
| 2012 | 6.84 | 7.74 | 0.035 | 4.87 | 5.34 | 0.021 |
| 2013 | 6.92 | 7.35 | 0.016 | 4.71 | 4.82 | 0.005 |
| 2014 | 5.88 | 6.9 | 0.042 | 4.4 | 4.78 | 0.018 |
| 2015 | 5.34 | 6.04 | 0.03 | 4.59 | 5.03 | 0.02 |
| 2016 | 4.94 | 5.51 | 0.025 | 4.67 | 5.17 | 0.023 |
| 2017 | 4.85 | 5.16 | 0.015 | 4.8 | 5.28 | 0.022 |
| 2018 | 5.06 | 5.33 | 0.012 | 4.78 | 5.11 | 0.016 |
| 2019 | 4.49 | 4.82 | 0.016 | 4.44 | 4.89 | 0.022 |
| 2020 | 3.41 | 4.13 | 0.038 | 4.33 | 4.65 | 0.016 |
| 2021 | 0 | 0.01 | 0.008 | 0.11 | 0.18 | 0.017 |
| Index month |  |  |  |  |  |  |
| 1 | 9.89 | 9.9 | 0 | 9.05 | 9.15 | 0.004 |
| 2 | 7.49 | 10.96 | 0.12 | 7.78 | 8.34 | 0.021 |
| 3 | 6.04 | 14.68 | 0.287 | 7.85 | 9.22 | 0.049 |
| 4 | 7.41 | 8.39 | 0.036 | 7.42 | 7.7 | 0.011 |
| 5 | 8.49 | 7.01 | -0.055 | 8.24 | 7.96 | -0.01 |
| 6 | 8.45 | 6.69 | -0.066 | 8.08 | 7.79 | -0.011 |
| 7 | 8.93 | 6.76 | -0.081 | 8.42 | 8.09 | -0.012 |
| 8 | 8.82 | 6.74 | -0.078 | 8.37 | 8 | -0.013 |
| 9 | 8.73 | 6.99 | -0.065 | 8.52 | 8.18 | -0.012 |
| 10 | 8.77 | 7.31 | -0.054 | 8.79 | 8.47 | -0.012 |
| 11 | 8.53 | 7.13 | -0.052 | 8.7 | 8.4 | -0.011 |
| 12 | 8.45 | 7.42 | -0.038 | 8.79 | 8.7 | -0.003 |
| Age (years) |  |  |  |  |  |  |
| Mean | 31.22 | 27.25 | -0.496 | 30.93 | 27.93 | -0.357 |
| Std. deviation | 4.68 | 6.49 |  | 4.88 | 6.82 |  |
| Median | 31 | 27 |  | 31 | 28 |  |
| Prior observation time (days) |  |  |  |  |  |  |
| Mean | 738.68 | 836.67 | 0.079 | 715.47 | 779.41 | 0.055 |
| Std. deviation | 744.33 | 985.54 |  | 741.19 | 894.21 |  |
| Median | 505 | 487 |  | 476 | 473 |  |
| Post observation time (days) |  |  |  |  |  |  |
| Mean | 1361.93 | 941.77 | -0.282 | 1221.32 | 929.25 | -0.213 |
| Std. deviation | 1196.94 | 883.18 |  | 1092.65 | 828.35 |  |
| Median | 932 | 640 |  | 825 | 641 |  |
| Pregnancy episode length (days) |  |  |  |  |  |  |
| Mean | 273.08 | 269.87 | -0.107 | 272.69 | 270.43 | -0.076 |
| Std. deviation | 18.59 | 23.46 |  | 18.99 | 23.11 |  |
| Median | 278 | 277 |  | 278 | 278 |  |
| Distinct conditions |  |  |  |  |  |  |
| Mean | 6.67 | 6.79 | 0.012 | 6.94 | 7.45 | 0.046 |
| Std. deviation | 6.72 | 7.37 |  | 7.38 | 8.18 |  |
| Median | 5 | 4 |  | 5 | 5 |  |
| Distinct drug ingredients |  |  |  |  |  |  |
| Mean | 4.68 | 4.62 | -0.007 | 4.14 | 4.45 | 0.041 |
| Std. deviation | 5.39 | 5.5 |  | 5.17 | 5.5 |  |
| Median | 3 | 3 |  | 3 | 3 |  |
| Distinct procedures |  |  |  |  |  |  |
| Mean | 7.39 | 6.61 | -0.075 | 7.07 | 6.92 | -0.014 |
| Std. deviation | 7.39 | 7.29 |  | 7.61 | 7.71 |  |
| Median | 5 | 5 |  | 5 | 5 |  |
| Distinct measurements |  |  |  |  |  |  |
| Mean | 6.69 | 6.3 | -0.023 | 13.61 | 12.48 | -0.03 |
| Std. deviation | 11.9 | 11.9 |  | 27.48 | 25.89 |  |
| Median | 3 | 3 |  | 3 | 3 |  |
| Distinct visit types |  |  |  |  |  |  |
| Outpatient Visit |  |  |  |  |  |  |
| Mean | 6.61 | 5.49 | -0.087 | 6.38 | 5.89 | -0.035 |
| Std. deviation | 9.81 | 8.3 |  | 10.48 | 8.98 |  |
| Median | 4 | 3 |  | 3 | 3 |  |
| Inpatient Visit |  |  |  |  |  |  |
| Mean | 0.11 | 0.11 | -0.001 | 0.1 | 0.11 | 0.003 |
| Std. deviation | 1.11 | 1.27 |  | 0.77 | 0.94 |  |
| Median | 0 | 0 |  | 0 | 0 |  |
| Emergency Room Visit |  |  |  |  |  |  |
| Mean | 0.16 | 0.34 | 0.092 | 0.38 | 0.33 | -0.007 |
| Std. deviation | 1.17 | 1.56 |  | 6.15 | 4.89 |  |
| Median | 0 | 0 |  | 0 | 0 |  |
| Charlson comorbidity index |  |  |  |  |  |  |
| Mean | 0.19 | 0.2 | 0.003 | 0.22 | 0.24 | 0.017 |
| Std. deviation | 0.85 | 0.89 |  | 0.93 | 1.02 |  |
| Median | 0 | 0 |  | 0 | 0 |  |

CCAE: IBM® MarketScan® Commercial Database; Clinformatics®: Optum's de-identified Clinformatics® Data Mart Database; Std.Diff.: Standardized difference of means

**Table E2.** Selected characteristics and standardized differences of linked and non-linked mothers in IBM® Marketscan® Commercial Database and Optum’s de-identified Clinformatics® Data Mart Database. Mother characteristics were measured on the pregnancy episode end date (index year and month, age) or during the 365-day period before and including the pregnancy episode end date (distinct event occurrence counts). Sensitivity algorithm implementation 2: all births, ±90d pregnancy episode end/infant date-of-birth correspondence.

| Characteristic | CCAE |  |  | Clinformatics® |  |  |
| --- | --- | --- | --- | --- | --- | --- |
|  | Linked<br>% (n = 2,567,501) | Non-linked<br>% (n = 965,224) | Std.Diff | Linked<br>% (n = 1,592,785) | Non-linked<br>% (n = 417,842) | Std.Diff |
| Index year |  |  |  |  |  |  |
| 2000 | 0.1 | 0.31 | 0.048 | 0.01 | 0.02 | 0.01 |
| 2001 | 0.66 | 0.84 | 0.021 | 2.02 | 2.19 | 0.012 |
| 2002 | 1.18 | 1.43 | 0.022 | 4.27 | 4.28 | 0 |
| 2003 | 2.24 | 2.2 | -0.002 | 4.82 | 4.66 | -0.007 |
| 2004 | 3.35 | 2.77 | -0.034 | 4.78 | 4.56 | -0.011 |
| 2005 | 4.11 | 3.35 | -0.04 | 5.07 | 4.04 | -0.05 |
| 2006 | 4.05 | 3.29 | -0.04 | 6.37 | 4.42 | -0.086 |
| 2007 | 4.84 | 4.14 | -0.034 | 6.3 | 4.84 | -0.064 |
| 2008 | 5.28 | 4.67 | -0.028 | 6.26 | 4.79 | -0.064 |
| 2009 | 6.37 | 5.39 | -0.042 | 5.8 | 4.59 | -0.055 |
| 2010 | 6.18 | 6.26 | 0.003 | 4.8 | 5.63 | 0.037 |
| 2011 | 7.63 | 6.84 | -0.031 | 4.49 | 6.14 | 0.073 |
| 2012 | 8.01 | 9.02 | 0.036 | 4.4 | 6.46 | 0.091 |
| 2013 | 6.7 | 7 | 0.012 | 5.07 | 4.91 | -0.008 |
| 2014 | 6.93 | 7.48 | 0.021 | 4.55 | 4.75 | 0.009 |
| 2015 | 5.44 | 6.21 | 0.033 | 4.47 | 4.87 | 0.019 |
| 2016 | 5.43 | 6.07 | 0.027 | 4.73 | 5.12 | 0.018 |
| 2017 | 4.93 | 5.31 | 0.017 | 4.68 | 5.21 | 0.024 |
| 2018 | 4.89 | 5.24 | 0.016 | 4.8 | 5.23 | 0.02 |
| 2019 | 4.94 | 5.18 | 0.011 | 4.74 | 5.11 | 0.017 |
| 2020 | 4.42 | 4.71 | 0.014 | 4.34 | 4.77 | 0.021 |
| 2021 | 2.32 | 2.3 | -0.001 | 3.23 | 3.44 | 0.012 |
| Index month |  |  |  |  |  |  |
| 1 | 8.12 | 6.59 | -0.059 | 7.56 | 7.42 | -0.005 |
| 2 | 7.78 | 6.13 | -0.065 | 7.43 | 7.04 | -0.015 |
| 3 | 8.77 | 6.73 | -0.076 | 8.33 | 7.99 | -0.012 |
| 4 | 8.61 | 6.51 | -0.08 | 8.15 | 7.81 | -0.012 |
| 5 | 8.99 | 6.81 | -0.081 | 8.58 | 8.12 | -0.017 |
| 6 | 8.73 | 7.1 | -0.06 | 8.47 | 8.19 | -0.01 |
| 7 | 8.77 | 7.34 | -0.052 | 8.77 | 8.5 | -0.01 |
| 8 | 8.76 | 7.37 | -0.051 | 8.96 | 8.74 | -0.008 |
| 9 | 8.56 | 7.51 | -0.039 | 8.72 | 8.62 | -0.003 |
| 10 | 9.55 | 9.18 | -0.013 | 8.75 | 8.87 | 0.004 |
| 11 | 8.25 | 11.09 | 0.096 | 8.26 | 8.67 | 0.015 |
| 12 | 5.11 | 17.64 | 0.403 | 8.03 | 10.02 | 0.07 |
| Age (years) |  |  |  |  |  |  |
| Mean | 31.98 | 27.86 | -0.519 | 31.67 | 28.65 | -0.361 |
| Std. deviation | 4.68 | 6.43 |  | 4.88 | 6.81 |  |
| Median | 32 | 27 |  | 32 | 28 |  |
| Prior observation time (days) |  |  |  |  |  |  |
| Mean | 1011.76 | 1106.54 | 0.077 | 988.16 | 1049.84 | 0.053 |
| Std. deviation | 744 | 985.32 |  | 740.95 | 894.03 |  |
| Median | 778 | 756 |  | 749 | 744 |  |
| Post observation time (days) |  |  |  |  |  |  |
| Mean | 1088.85 | 671.9 | -0.281 | 948.63 | 658.82 | -0.212 |
| Std. deviation | 1195.92 | 882.7 |  | 1091.6 | 827.58 |  |
| Median | 659 | 370 |  | 553 | 370 |  |
| Distinct conditions |  |  |  |  |  |  |
| Mean | 19.78 | 20.82 | 0.068 | 20.19 | 21.44 | 0.078 |
| Std. deviation | 10.21 | 11.43 |  | 10.65 | 11.96 |  |
| Median | 17 | 18 |  | 18 | 19 |  |
| Distinct drug ingredients |  |  |  |  |  |  |
| Mean | 5.29 | 5.67 | 0.049 | 5.4 | 6 | 0.075 |
| Std. deviation | 5.34 | 5.63 |  | 5.39 | 5.84 |  |
| Median | 4 | 4 |  | 4 | 4 |  |
| Distinct procedures |  |  |  |  |  |  |
| Mean | 17.44 | 16.84 | -0.05 | 18.59 | 18.55 | -0.003 |
| Std. deviation | 8.44 | 8.52 |  | 8.72 | 9.1 |  |
| Median | 16 | 15 |  | 18 | 17 |  |
| Distinct measurements |  |  |  |  |  |  |
| Mean | 20.8 | 21.03 | 0.01 | 43.99 | 41.65 | -0.046 |
| Std. deviation | 15.54 | 16.65 |  | 36.44 | 36.04 |  |
| Median | 18 | 18 |  | 32 | 27 |  |
| Distinct visit types |  |  |  |  |  |  |
| Outpatient Visit |  |  |  |  |  |  |
| Mean | 15.52 | 14.2 | -0.089 | 15.7 | 14.76 | -0.054 |
| Std. deviation | 11.33 | 9.52 |  | 13.45 | 10.96 |  |
| Median | 13 | 12 |  | 12 | 12 |  |
| Inpatient Visit |  |  |  |  |  |  |
| Mean | 1.13 | 1.14 | 0.006 | 1.14 | 1.12 | -0.018 |
| Std. deviation | 0.68 | 0.69 |  | 0.72 | 0.64 |  |
| Median | 1 | 1 |  | 1 | 1 |  |
| Emergency Room Visit |  |  |  |  |  |  |
| Mean | 0.33 | 0.67 | 0.149 | 1.03 | 0.98 | -0.007 |
| Std. deviation | 1.37 | 1.85 |  | 5.63 | 4.52 |  |
| Median | 0 | 0 |  | 0 | 0 |  |
| Charlson comorbidity index |  |  |  |  |  |  |
| Mean | 0.3 | 0.3 | 0.001 | 0.32 | 0.34 | 0.016 |
| Std. deviation | 0.9 | 0.94 |  | 0.98 | 1.08 |  |
| Median | 0 | 0 |  | 0 | 0 |  |

CCAE: IBM® MarketScan® Commercial Database; Clinformatics®: Optum's de-identified Clinformatics® Data Mart Database; Std.Diff.: Standardized difference of means

**Table E3.** Selected characteristics and standardized differences of linked and non-linked infants in IBM® Marketscan® Commercial Database and Optum’s de-identified Clinformatics® Data Mart Database. Infant characteristics were measured on the inferred pregnancy episode end date (index year and month, age) or during the 365-day period after and including the inferred pregnancy episode end date. (Distinct event occurrence counts). Sensitivity algorithm implementation 2: all births, ±90d pregnancy episode end/infant date-of-birth correspondence

| Characteristic | CCAE |  |  | Clinformatics® |  |  |
| --- | --- | --- | --- | --- | --- | --- |
|  | Linked<br>% (n = 2,567,501) | Non-linked<br>% (n = 2,367,877) | Std.Diff | Linked<br>% (n = 1,592,785) | Non-linked<br>% (n = 1,787,026) | Std.Diff |
| Sex |  |  |  |  |  |  |
| gender = FEMALE | 48.72 | 48.65 | -0.002 | 48.58 | 48.55 | -0.001 |
| gender = MALE | 51.28 | 51.35 | 0.002 | 51.42 | 51.45 | 0.001 |
| Index year |  |  |  |  |  |  |
| 2000 | 0.1 | 1.1 | 0.131 | 0.01 | 3.88 | 0.283 |
| 2001 | 0.66 | 1.06 | 0.043 | 2.02 | 6.9 | 0.238 |
| 2002 | 1.18 | 2.52 | 0.1 | 4.27 | 4.93 | 0.031 |
| 2003 | 2.24 | 3.95 | 0.099 | 4.82 | 4.65 | -0.008 |
| 2004 | 3.35 | 4.28 | 0.048 | 4.78 | 4.48 | -0.014 |
| 2005 | 4.11 | 4.42 | 0.015 | 5.07 | 6.65 | 0.067 |
| 2006 | 4.05 | 4.98 | 0.045 | 6.37 | 6.07 | -0.013 |
| 2007 | 4.84 | 4.77 | -0.003 | 6.3 | 5.97 | -0.014 |
| 2008 | 5.28 | 6.23 | 0.041 | 6.26 | 5.06 | -0.052 |
| 2009 | 6.37 | 6.58 | 0.009 | 5.8 | 4.13 | -0.077 |
| 2010 | 6.18 | 7.52 | 0.053 | 4.8 | 3.56 | -0.062 |
| 2011 | 7.63 | 7.38 | -0.01 | 4.49 | 3.51 | -0.05 |
| 2012 | 8.01 | 7.27 | -0.028 | 4.4 | 3.51 | -0.046 |
| 2013 | 6.7 | 6.33 | -0.015 | 5.07 | 3.99 | -0.052 |
| 2014 | 6.93 | 6.32 | -0.025 | 4.55 | 4.02 | -0.026 |
| 2015 | 5.44 | 4.14 | -0.061 | 4.47 | 4.68 | 0.01 |
| 2016 | 5.43 | 4.08 | -0.064 | 4.73 | 4.84 | 0.005 |
| 2017 | 4.93 | 4.02 | -0.044 | 4.68 | 4.68 | 0 |
| 2018 | 4.89 | 4.49 | -0.019 | 4.8 | 4.44 | -0.017 |
| 2019 | 4.94 | 3.66 | -0.064 | 4.74 | 4.04 | -0.034 |
| 2020 | 4.42 | 3.43 | -0.051 | 4.34 | 3.49 | -0.044 |
| 2021 | 2.32 | 1.49 | -0.061 | 3.23 | 2.54 | -0.041 |
| Index month |  |  |  |  |  |  |
| 1 | 8.12 | 4.63 | -0.143 | 7.56 | 6.16 | -0.055 |
| 2 | 7.78 | 6.46 | -0.051 | 7.43 | 5.92 | -0.06 |
| 3 | 8.77 | 7.53 | -0.045 | 8.33 | 6.73 | -0.061 |
| 4 | 8.61 | 8.45 | -0.006 | 8.15 | 6.96 | -0.045 |
| 5 | 8.99 | 8.57 | -0.015 | 8.58 | 10.16 | 0.054 |
| 6 | 8.73 | 9.02 | 0.01 | 8.47 | 8.49 | 0.001 |
| 7 | 8.77 | 10.49 | 0.059 | 8.77 | 9.93 | 0.04 |
| 8 | 8.76 | 9.87 | 0.038 | 8.96 | 9.57 | 0.021 |
| 9 | 8.56 | 10.3 | 0.06 | 8.72 | 9.95 | 0.042 |
| 10 | 9.55 | 9.62 | 0.002 | 8.75 | 9.38 | 0.022 |
| 11 | 8.25 | 7.84 | -0.015 | 8.26 | 8.32 | 0.002 |
| 12 | 5.11 | 7.22 | 0.088 | 8.03 | 8.43 | 0.015 |
| Post observation time (days) |  |  |  |  |  |  |
| Mean | 448.1 | 879.59 | 0.27 | 599.91 | 750.73 | 0.101 |
| Std. deviation | 1154.43 | 1107.74 |  | 1127.43 | 985.22 |  |
| Median | 0 | 484 |  | 118 | 395 |  |
| Distinct conditions |  |  |  |  |  |  |
| Mean | 6.69 | 5.74 | -0.101 | 7.73 | 6.39 | -0.129 |
| Std. deviation | 6.87 | 6.4 |  | 7.64 | 7.1 |  |
| Median | 5 | 4 |  | 6 | 5 |  |
| Distinct drug ingredients |  |  |  |  |  |  |
| Mean | 2.63 | 2.45 | -0.038 | 8.53 | 7.44 | -0.065 |
| Std. deviation | 3.38 | 3.38 |  | 12.02 | 11.76 |  |
| Median | 1 | 1 |  | 2 | 2 |  |
| Procedures |  |  |  |  |  |  |
| Mean | 12.74 | 9.9 | -0.279 | 12.25 | 9.99 | -0.21 |
| Std. deviation | 7.38 | 7.01 |  | 7.82 | 7.36 |  |
| Median | 12 | 9 |  | 11 | 9 |  |
| Distinct measurements |  |  |  |  |  |  |
| Mean | 2.84 | 2.24 | -0.086 | 3.28 | 2.83 | -0.042 |
| Std. deviation | 5.05 | 4.93 |  | 7.41 | 7.61 |  |
| Median | 1 | 1 |  | 1 | 1 |  |
| Distinct visit types |  |  |  |  |  |  |
| Outpatient Visit |  |  |  |  |  |  |
| Mean | 10.34 | 8.66 | -0.182 | 9.45 | 8.32 | -0.105 |
| Std. deviation | 6.59 | 6.47 |  | 7.72 | 7.5 |  |
| Median | 9 | 8 |  | 8 | 7 |  |
| Inpatient Visit |  |  |  |  |  |  |
| Mean | 0.98 | 0.5 | -0.648 | 1.03 | 0.69 | -0.491 |
| Std. deviation | 0.47 | 0.57 |  | 0.47 | 0.51 |  |
| Median | 1 | 0 |  | 1 | 1 |  |
| Emergency Room Visit |  |  |  |  |  |  |
| Mean | 0.22 | 0.24 | 0.016 | 0.1 | 0.13 | 0.006 |
| Std. deviation | 0.84 | 0.91 |  | 3.94 | 3.45 |  |
| Median | 0 | 0 |  | 0 | 0 |  |

CCAE: IBM® MarketScan® Commercial Database; Clinformatics®: Optum's de-identified Clinformatics® Data Mart Database; Std.Diff.: Standardized difference of means

## Footnotes

1 Charlson, M.E., et al., A new method of classifying prognostic comorbidity in longitudinal studies: development and validation. J Chronic Dis, 1987. **40**(5): p. 373-83.

